# Biopsychosocial correlates of somatic symptom burden in chronic kidney disease: results of the Hamburg City Health Study (HCHS)

**DOI:** 10.1101/2025.11.27.25341140

**Authors:** Birte Jessen, Bernd Löwe, Raphael Twerenbold, Martin Härter, Volker Harth, Hanno Hoven, Christian Schmidt-Lauber, Tobias B. Huber, Meike Shedden-Mora

## Abstract

**Background:** Persistent somatic symptoms (PSS) in individuals with chronic kidney disease (CKD) occur across all stages and impact patients’ quality of life, morbidity and mortality. We aimed to unravel associations between biopsychosocial factors and symptom burden in individuals with CKD.

**Methods:** This cross-sectional study analysed individuals with CKD (estimated glomerular filtration rate (eGFR) <60 mL/min/1.73m^2^) from the first 10,000 participants of the population-based Hamburg City Health Study (HCHS). Somatic symptom burden (PHQ-15) was the primary outcome. Its association with potential biological (e.g., eGFR), psychological (e.g., depressive symptoms), and sociodemographic correlates was analysed in a multivariate prediction model. Correlates were compared to healthy controls and individuals with coronary heart disease (CHD).

**Results:** Somatic symptom burden in patients with non-dialysis CKD (n = 582, mean age: 69.58 years; 44.7% women; mean baseline eGFR: 52 mL/min/1.73m^2^) increased with lower eGFR (r = -.161, *p* < .001). However, in a stepwise multiple linear regression model, eGFR was not correlated with symptom burden. In contrast, female sex, coronary heart disease, self-reported general health, proneness to illness, and depressive symptoms were associated with somatic symptom burden. Correlates of somatic symptom burden in patients with CHD (n = 618, mean age: 67.13 years; 24.1% women; mean baseline eGFR: 81 mL/min/1.73m^2^) and matched healthy controls (n = 582, mean age: 69.58 years; 44.7% women; mean baseline eGFR: 81 mL/min/1.73m^2^) were comparable.

**Conclusion:** Somatic symptom burden in CKD was only marginally related to the eGFR but rather to biopsychosocial factors. Therefore, taking a biopsychosocial perspective on PSS in CKD is important.

**Key points:**

- Kidney function (GFR) correlated with somatic symptom burden bivariately, but not in the multiple regression model.
- Female sex, coronary heart disease, self-reported general health, proneness to illness and depressive symptoms correlated to symptom burden.
- Similar biopsychosocial correlates of symptom burden were observed in individuals with CKD, coronary heart disease (CHD) and controls.

## Introduction

Persistent somatic symptoms (PSS) describe subjectively bothersome somatic complaints that are present on most days for at least several months, regardless of their cause.^1,2^ PSS occur in approximately 15% of the general population^3^, and in most chronic diseases.^1–4^ PSS in chronic kidney disease (CKD) appear in all stages and are associated with quality of life (QoL), morbidity and mortality.^5,6^ Recent evidence highlights that even individuals with early CKD report a high somatic symptom burden with most prevalent symptoms being fatigue (70%), pruritus (46%) and bone or joint pain (55%).^7,8^

Contrary to clinical expectations, scientific evidence if symptom burden severity is related to CKD severity remains inconclusive. Reported correlations between the estimated glomerular filtration rate (eGFR) and general subjective symptom burden are weak^5,8^ and overall symptom burden^4,7^ is comparable across non-dialysis and end-stage kidney disease.^5,9^ In contrast to a comparable overall symptom burden however, specific symptoms, namely lack of appetite, fatigue, impaired sex life, and cramps were recently found to increase with CKD severity.^7^ Symptom burden in CKD seems to be comparable to that of other chronic diseases such as coronary heart disease (CHD)^10^, or even terminal malignant conditions.^11^ In fact, patients often report that disabling somatic symptoms are the primary disease burden in their lives.^12^

Although the etiology of PSS in CKD is poorly understood and researched, a stronger focus on disease-overarching biopsychosocial factors contributing to PSS across diseases has recently been proposed, emphasizing a biopsychosocial perspective.^1,2,4^ From this perspective, psychological and health-related factors such as subjective illness beliefs, expectations, depression, anxiety, cognitive-perceptual mechanisms, emotion regulation strategies, and adverse childhood experiences are thought to contribute to somatic symptom burden.^13,14^ In CKD, initial evidence highlights the importance of depression and anxiety, which were either associated with elevated somatic symptom burden and adverse outcomes^15–17^ or higher compared to controls.^18,19^ Adverse childhood experiences were identified as major predictor of symptom burden and mental and physical health^20,21^ and were associated with higher all-cause mortality in individuals with CKD.^18,22^ Other factors identified as predictors of non-adherence, depression, morbidity, mortality, and lower QoL in CKD include patients’ illness perceptions^19^ and physical inactivity.^23–25^

While our knowledge on biopsychosocial correlates of PSS in CKD is still limited, accumulating evidence suggests pathophysiological mechanisms of PSS in CKD being comparable to those in somatoform and functional disorders.^26^ The development of PSS in CKD is probably rather driven by the complex interplay of biomedical (comorbidities), treatment-related (medication side effects, dialysis), and psychological factors (depression, anxiety).^27^

The primary study aim was to analyse cross-sectional sociodemographic, biological, psychological and health-related correlates of somatic symptom burden in individuals with CKD. We hypothesized that psychosocial factors (depression, anxiety) are stronger associated with somatic symptom burden in individuals with CKD than biomedical factors (eGFR, medication). As secondary outcome, we analysed biopsychosocial correlates of QoL. Third, we aimed to compare the impact of biopsychosocial correlates on symptom burden in CKD with a healthy and a somatic control group with coronary heart disease (CHD). Comparing CKD and CHD may provide important insights into disease-specific and disease-overarching mechanisms. We expected psychosocial correlates of somatic symptom burden to be comparable in individuals with CKD and matched somatic and healthy controls.

## Materials and Methods

### Study design and population

Data was taken from the Hamburg City Health Study (HCHS), a prospective, long-term, population-based cohort study designed to discover important risk and prognostic factors in major chronic diseases.^28^ The HCHS includes individuals between 45 and 74 years of age recruited from the general population of Hamburg, Germany via mail. Cross-sectional data from the first 10,000 participants, collected between February 2016 and November 2018, were available for this analysis. Based on this cohort, we analysed three different groups in this cross-sectional study. First, individuals with CKD, defined by an eGFR <60ml/min/1.73m² were identified.^29^ The eGFR was calculated with the 2009 Chronic Kidney Disease Epidemiology Collaboration (CKD-Epi) formula for creatinine.^29,30^ Creatinine measurements were performed as described recently^31^, and eGFR was staged according to recommendations of the Kidney Disease: Improving Global Outcomes (KDIGO) Collaborative.^29^ Only a comorbid coronary heart disease was not excluded in the CKD group. Second, a control group with coronary heart disease (CHD) and an eGFR >60ml/min/1.73m^2^ was defined to compare correlates of somatic symptom burden to a comparable chronic illness. A certain overlap or simultaneous presence of CKD and CHD should be mentioned, but these subjects were categorised in the CKD group. Third, a healthy control sample with neither CHD nor CKD matched in sex, age and education status with no tolerance for deviations and furthermore no distinctions regarding age, sex and educational status to the CKD-cohort using case-control-matching via SPSS was included. Later, demographic and clinical variables across CKD stages were compared. Stages ≥ 3b were combined due to small number of participants. The HCHS study protocol was approved by the ethics committee of the State of Hamburg Chamber of Medical Practitioners (PV5131) and was registered at ClinicalTrial.gov (NCT03934957). The analysis was preregistered at the Open Science Framework (https://osf.io/58s2r).

#### Assessment and study outcomes

*Primary Outcome: **Somatic symptom burden***

*Somatic symptom burden* was measured using the Patient-Health-Questionnaire-15 (PHQ-15), a self-report questionnaire measuring the severity of 15 symptomatic symptoms (see S2) on a three-point scale ranging from 0 (*not bothered at all)* to 2 (*bothered a lot).* Scores range from 0 to 30, categorized as *low* (0-4 points), *medium* (5-9 points) and *high* (≥10 points).^32^ Internal consistency and test-retest reliability and a good fit for large-scale studies of the score has been shown in different studies.^33^

*Secondary Outcome: **Quality of Life***

QoL was measured using the European Quality of Life 5 Dimensions 5 Level Version (EQ5D-5L), a standardised 6-item self-report questionnaire with five dimensions (*mobility*, *self-care*, *usual activities*, *pain/discomfort* & *anxiety/depression*) rated on a 5-point scale. The sixth item represents a visual analogue scale from 0-100 rating subjectively perceived general health status.^34^

#### Biopsychosocial correlates

*Sociodemographic factors* included age, sex and education status staged according to the International Standard Classification of Education (ISCED 2011) of *low* (Levels 0-2), *medium* (Levels 3-4), *high* (Levels 5-8).^35^ Vitals and non-renal biomarkers such as high sensitive C-reactive protein (hsCRP) were measured as described.^31,36^

*Comorbidities* were assessed using composite scores, for coronary heart disease (self-reported diagnosis of coronary artery disease, myocardial infarction, balloon dilatation of the coronary arteries (PTCA), stent implantation or bypass surgery), arterial hypertension (antihypertensive medication, systolic blood pressure >140mmHg, diastolic blood pressure >90mmHg or self-report), diabetes mellitus (self-report, antidiabetic medication, fasting glucose >126mg/dl, non-fasting glucose >200mg/dl), and dyslipidaemia (LDL/HDL ratio >3.5 or lipid modifying drugs). Medication^28^ and the presence of further chronic illnesses (asthma bronchiale, chronic obstructive bronchitis and cancer) were assessed via self-report.

*Health related factors* were measured with variables from the DEGS study (study on the health of adults in Germany). General health, proneness to illness and the expectancy of deterioration of health states were rated on a five-point scale (e.g. general health: 1 *excellent* to 5 *bad*). Physician visits in the past year were assessed via self-report.^37^

*Smoking behaviour* was assessed via self-report and dichotomized into *yes* and *no* (combination of non-smokers and former smokers).

*Physical activity* was measured with single items from a spaq (seasonal pattern assessment questionnaire) based on the *European Prospective Investigation into Cancer and Nutrition (EPIC)* which was developed by the DIFE (German Institute of Human Nutrition) assessing physical activity in hours per week.^38^

*Psychological factors*: Depressive symptoms were assessed with the *Patient-Health-Questionnaire 9 (PHQ-9),* a valid, reliable self-report questionnaire consisting of 9 items on a four-point scale (0 *not at all* to 3 *nearly every day)* categorized as *mild* (10-14), *medium* (15-29) and *high* (20-27).^39^

*Anxiety* was assessed with the *Generalized-Anxiety-Disorder-7 (GAD-7)*, a valid, reliable self-report 7-item questionnaire assessing symptoms on a four-point scale (0 *not at all* to 3 *nearly every day)* and cut-off points classifying *mild* (5-9), *moderate* (10-14) and *severe* (15-21) anxiety.^40^

*Adverse Childhood Experiences* were assessed using the same named 10-item questionnaire (ACE) for five types of childhood traumatization (e.g. *sexual* and *physical abuse*) with a dichotomous response format (no/yes) and a sum score by summing all ACEs.^41^

### Data analysis

Statistical analyses were carried out using SPSS version 27. Missing data in the self-report scales (PHQ-15, PHQ-9, GAD7 and EQ5D), ranging from 7.6 to 12.8% was imputed using the expectation-maximization-algorithm if at least one item was completed. Missing educational status variables (5.8%) were replaced with a medium education level. Missing data in the other variables ranged from 3.1% (haemoglobin) to 33.6% (expectation of deterioration) which were not imputed. Because of similar group sizes of the CKD and CHD group, only the healthy controls were matched.

To analyse biopsychosocial correlates of somatic symptom burden in CKD a stepwise multiple linear regression model with four steps was conducted. Variables were included in the model in the following order: 1) sociodemographic factors (age, sex and education), 2) biomedical factors (eGFR, body mass index (BMI), coronary heart disease, arterial hypertension, diabetes mellitus, dyslipidaemia and medication), 3) health-related factors (physical activity, smoking behaviour, general health status, proneness to illness, expectancy of deterioration and sum of physician visits per year), 4) psychological factors (depressive symptoms, anxiety and adverse childhood experiences). The same regression model was applied to examine the influence of these factors on QoL in CKD. Analogous regression models were calculated in the CHD and the healthy control group to compare correlates across these groups. Odds ratios were calculated with logistic regression models to compare the existence of single symptoms of the PHQ-15 in all groups. To examine potential differences of biopsychosocial correlates of somatic symptom burden within the CKD group, ANOVAs and Chi-Square tests were then calculated.

## Results

### Sample characteristics

The CKD cohort included 582 individuals and 618 individuals had coronary heart disease. The healthy control group constituted of 582 individuals. The study population was predominantly white. Table 1 provides an overview of the demographic and clinical characteristics of the overall HCHS cohort and the three subgroups. Participants with CKD and the healthy control sample were significantly older than participants with CHD (*F* = 28.12, *p* < .001). The percentage of women was higher in the CKD and healthy control group compared to the CHD group (χ² = 72.80, *p* < .001). Scores of the psychological assessments were higher in the CHD group for general symptom burden (*F* = 9.75, *p* < .001), depressive symptoms (*F* = 8.16, *p* < .001), and adverse childhood experiences (*F* = 4.85, *p* = .008) when comparing to healthy controls. Anxiety scores were the highest in the CHD group (*F* = 4.86, *p* = .008). In turn, the healthy control group scored highest on the QoL scale (*F* = 14.65, *p* < .001).

**Table 1:** Demographic and clinical characteristics of participants.

| Characteristic | cohort of 10.000 | CKD cohort<sup>a</sup> | CHD control group<sup>b</sup> | healthy control group<sup>c</sup> | comparison |
| --- | --- | --- | --- | --- | --- |
| n | 10.000 | 582 | 618 | 582 |  |
| **Sociodemographic variables** |  |  |  |  |  |
| Age mean ± SD; (range) | 62.37 ± 8.45 (46-78) | 69.58 ± 6.37 (47-78) | 67.13 ± 6.92 (47-78) | 69.58 ± 6.37 (47-78) | $t = 6.39, p < .001 (a > b)$ |
| Sex, female n (%) | 5108 (51.1%) | 260 (44.7%) | 149 (24.1%) | 260 (44.7%) | $\chi^2 = 56.41, p < .001 (a > b)$ |
| Education n (%) |  |  |  |  |  |
| Low | 466 (4.7%) | 42 (7.2.0%) | 35 (5.7%) | 42 (7.2%) |  |
| Medium | 4721 (47.2%) | 293 (50.3%) | 293 (47.4%) | 298 (51.2%) |  |
| High | 4232 (42.3%) | 196 (33.7%) | 250 (40.5%) | 196 (33.7%) | $\chi^2 = 5.19, p = .08$ |
| Ethnicity |  |  |  |  | - |
| White | 9802 (98.0%) | 572 (98.3%) | 611 (98.9%) | 573 (98.5%) |  |
| Hispanic | 35 (0.4%) | 2 (0.3%) | 1 (0.2%) | 1 (0.2%) |  |
| Asian | 57 (0.6%) | 0 (0.0%) | 1 (0.2%) | 3 (0.5%) |  |
| Black | 36 (0.4%) | 1 (0.2%) | 1 (0.2%) | 2 (0.3%) |  |
| Other | 40 (0.4%) | 2 (0.3%) | 3 (0.5%) | 2 (0.3%) |  |
| **Health-related assessments** |  |  |  |  |  |
| Physical activity in h/week mean ± SD; (range) | 2.61 ± 3.00 (0.00-20.00) | 2.30 ± 3.34 (0.00-20.00) | 2.55 ± 3.11 (0.00-20.00) | 2.76 ± 3.34 (0.00-20.00) | $F = 2.44, p = .09$ |
| Current smoker n (%) | 1978 (19.8%) | 85 (14.6%) | 113 (18.3%) | 86 (14.8%) | $\chi^2 = 3.92, p = .141$ |
| eGFR mean ± SD; (range) | 84 ± 14 (6-127) | 52 ± 8 (7-59) | 81 ± 11 (60-111) | 81 ± 11 (60-112) | $F = 1627.23, p = .000 (a < b, c)$ |
| Haemoglobin (g/dl) mean ± SD; (range) | 14.34 ± 1.14 (8.3-18.9) | 14.12 ± 1.39 (9.5-18.7) | 14.60 ± 1.20 (9.9-18.0) | 14.43 ± 1.19 (9.0-18.1) | $F = 21.55, p < .001 (a < b, c)$ |
| hs CRP (mg/dl) mean ± SD; (range) | 0.24 ± 0.44 (0.01-11.70) | 0.31 ± 0.46 (0.01-6.25) | 0.24 ± 0.38 (0.01-4.35) | 0.27 ± 0.47 (0.01-5.47) | $F = 3.90, p = .02 (a > b)$ |
| BMI (kg/m<sup>2</sup>) mean ± SD; (range) | 26.76 ± 4.69 (15.59-61.63) | 28.75 ± 5.34 (18.31-48.51) | 28.06 ± 4.61 (16.84-61.63) | 26.94 ± 4.44 (16.91-45.59) | $F = 19.90, p < .001 (a, b > c; b > c)$ |
| Systolic blood pressure (mmHg) mean ± SD; (range) | 138.84 ± 19.36 (65.0-249.5) | 143.13 ± 21.14 (92.0-249.5) | 141.20 ± 18.25 (87.5-206.5) | 144.48 ± 20.79 (65.0-223.0) | $F = 3.90, p = .02 (b < c)$ |
| Diastolic blood pressure (mmHg) mean ± SD; (range) | 82.09 ± 10.04 (41.5-131.0) | 80.58 ± 10.96 (41.5-125.0) | 80.71 ± 10.53 (49.5-112.5) | 81.82 ± 10.41 (50.0-119.0) | $F = 2.30, p = .101$ |
| Heart frequency (bpm) mean ± SD; (range) | 69.70 ± 11.10 (29.0-129.0) | 70.01 ± 12.02 (34.0-119.5) | 68.12 ± 11.55 (34.0-107.0) | 70.45 ± 11.77 (35.0-117.0) | $F = 6.39, p = .002 (a > b, b < c)$ |
| Current medication n (%) | 7539 (75.4%) | 508 (87.3%) | 579 (93.7%) | 470 (80.8%) | $\chi^2 = 51.45, p < .001 (a < b; a, b > c)$ |
| Diabetes mellitus n (%) | 794 (7.9%) | 126 (21.6%) | 99 (16.0%) | 59 (10.1%) | $\chi^2 = 28.17, p < .001 (a > b, c; b > c)$ |
| Coronary heart disease n (%) | 758 (7.6%) | 109 (18.7%) | 618 (100%) | - | $\chi^2 = 1414.44, p < .001 (a < b)$ |
| Arterial hypertension n (%) | 6301 (66.0%) | 507 (87.1%) | 568 (91.9%) | 435 (74.7%) | $\chi^2 = 72.74, p < .001 (a < b; a, b > c)$ |
| Dyslipidaemia n (%) | 2276 (22.8%) | 252 (43.3%) | 432 (69.9%) | 159 (27.3%) | $\chi^2 = 218.17, p < .001 (a < b; a, b > c)$ |
| Asthma bronchiale n (%) | 861 (8.6%) | 47 (8.1%) | 54 (8.7%) | 51 (8.8%) | $\chi^2 = 0.16, p = .92$ |
| Chronic obstructive bronchitis n (%) | 629 (6.3%) | 44 (7.6%) | 65 (10.5%) | 39 (6.7%) | $\chi^2 = 5.90, p = .05$ |
| Cancer n (%) | 1592 (15.9%) | 158 (27.1%) | 115 (18.6%) | 113 (19.4%) | $\chi^2 = 16.20, p < .001 (a > b, c)$ |
| **Psychological variables** |  |  |  |  |  |
| Somatic symptom burden (PHQ-15) mean ± SD | 4.99 ± 3.99 | 5.63 ± 4.23 | 6.20 ± 4.71 | 5.04 ± 4.10 | $F = 9.75, p < .001 (b > c)$ |
| Low symptom burden (0-4) n (%) | 4994 (49.9%) | 233 (40.0%) | 246 (39.8%) | 295 (50.7%) |  |
| Medium symptom burden (5-9) n (%) | 2972 (29.7%) | 185 (31.8%) | 180 (29.1%) | 170 (29.2%) |  |
| High symptom burden ($\geq 10$) n (%) | 1212 (12.1%) | 95 (16.3%) | 136 (22.0%) | 66 (11.3%) | $\chi^2 = 31.23, p < .001 (a, c < b)$ |
| Depressive symptoms (PHQ-9) mean ± SD | 3.54 ± 3.56 | 3.52 ± 3.49 | 4.01 ± 3.95 | 3.12 ± 3.47 | $F = 8.16, p < .001 (b > c)$ |
| Anxiety (GAD7) mean ± SD | 2.78 ± 3.04 | 2.47 ± 2.90 | 2.94 ± 3.22 | 2.43 ± 2.86 | $F = 4.86, p = .008 (a, c < b)$ |
| Adverse Childhood Experience (ACE) mean ± SD | 1.16 ± 1.63 | 1.10 ± 1.53 | 1.22 ± 1.75 | 0.92 ± 1.43 | $F = 4.85, p = .008 (b > c)$ |
| Quality of life (EQ5D-5L) mean ± SD | 0.91 ± 0.13 | 0.87 ± 0.16 | 0.88 ± 0.17 | 0.92 ± 0.11 | $F = 14.65, p < .001 (a, b < c)$ |
| Visual analogue scale of quality of life (EQ5D-5L) mean ± SD | 79.19 ± 15.26 | 72.80 ± 18.52 | 72.20 ± 18.85 | 79.02 ± 14.39 | $F = 25.33, p < .001 (a, b < c)$ |

Note. Sample sizes across variables may differ due to missing values. Comparisons were analysed using Bonferroni post hoc tests. Bold types stand for significant values.
CKD, chronic kidney disease; CHD, coronary heart disease; eGFR, estimated glomerular filtration rate; hsCRP, highly sensitive C-reactive protein; BMI, Body-Mass-Index; PHQ-15, Patient-Health-Questionnaire-15; PHQ-9, Patient-Health-Questionnaire-9; GAD-7, Generalized-Anxiety-Disorder-7; ACE, Adverse Childhood Experiences; EQ5D-5L, European Quality of Life 5 Dimensions 5 Level Version; SD, standard deviation; t, t-value; n, number; $\chi^2$ , chi square coefficient; p, p-value; F, F-value;

### Biopsychosocial correlates of somatic symptom burden

In the CKD group, lower eGFR (*r* = -.161, *p* < .001) and a higher BMI (*r* = .116, *p* = .01) were correlated with higher symptom burden. Depressive symptoms (*r* = .675, *p* < .001), anxiety (*r* = .543, *p* < .001), adverse childhood experiences (*r* = .271, *p* < .001), QoL (*r* = -.538, *p <* .001) as well as the sum of physician visits per year (*r* = .245, *p* < .001) correlated significantly with somatic symptom burden (see supplemental table S1 for all bivariate correlations with symptom burden and QoL).

Analysing biopsychosocial correlates of somatic symptom burden, a stepwise multiple linear regression model was calculated in the CKD sample (Table 2). Female sex was associated with higher somatic symptom burden (β = .143, *p* < .001). Only comorbid coronary heart disease as biological factor was associated with higher symptom burden (β = .113, *p* = .005), while eGFR was not (β = -.053, *p* = .15). Self-reported poor general health status (β = .201, *p* < .001) was an associated health-related factor. Depressive symptoms were associated with high somatic symptom burden (β = .469, *p* < .001). The final regression model explained 57.2% of the variance in somatic symptom burden.

**Table 2:** Correlates of somatic symptom burden in individuals with CKD, CHD and healthy controls using a stepwise multiple linear regression model.

| Steps | CKD group |  | CHD group |  | healthy control group |  |
| --- | --- | --- | --- | --- | --- | --- |
|  | Standardized beta | p-value | Standardized beta | p-value | Standardized beta | p-value |
| **Step 1: sociodemographic factors** | $(\vert R^2\vert = .071, p < .001; \Delta R^2 = .079; p < .001)$ | | $(\vert R^2\vert = .057, p < .001; \Delta R^2 = .064; p < .001)$ | | $(\vert R^2\vert = .019, p = .019; \Delta R^2 = .027; p = .019)$ | |
| Age | .018 | .63 | **.150** | **<.001** | .074 | .06 |
| Female sex | **.143** | **<.001** | **.129** | **<.001** | .065 | .09 |
| Education | -.031 | .41 | .005 | .89 | .000 | 1.00 |
| **Step 2: biological factors** | $(\vert R^2\vert = .147, p < .001; \Delta R^2 = .092; p < .001)$ | | $(\vert R^2\vert = .071, p = .060; \Delta R^2 = .027; p < .001)$ | | $(\vert R^2\vert = .065, p < .001; \Delta R^2 = .061; p < .001)$ | |
| eGFR | -.053 | .15 | -.012 | .75 | **.080** | **.03** |
| BMI | .000 | .99 | .051 | .16 | .056 | .14 |
| Coronary heart disease | **.113** | **.005** | - | - | - | - |
| Arterial hypertension | .009 | .82 | -.005 | .89 | .044 | .24 |
| Diabetes mellitus | .054 | .15 | -.032 | .36 | .043 | .24 |
| Dyslipidaemia | .005 | .91 | -.015 | .67 | .011 | .75 |
| Medication | -.052 | .16 | -.021 | .56 | .020 | .59 |
| **Step 3: health-related factors** | $(\vert R^2\vert = .374, p < .001; \Delta R^2 = .232; p < .001)$ | | $(\vert R^2\vert = .364, p < .001; \Delta R^2 = .295; p < .001)$ | | $(\vert R^2\vert = .316, p < .001; \Delta R^2 = .257; p < .001)$ | |
| Physical activity in h/week | -.007 | .86 | -.037 | .28 | -.028 | .44 |
| Smoking behavior | -.026 | .47 | .011 | .76 | -.049 | .18 |
| General state of health | **.201** | **<.001** | **.185** | **<.001** | **.117** | **.006** |
| Proneness to illness | -.065 | .11 | **-.076** | **.05** | **-.077** | **.04** |
| Expectancy of deterioration of health status | -.013 | .73 | -.025 | .48 | **-.120** | **.001** |
| Sum of physician visits per year | .070 | .07 | **.088** | **.01** | .039 | .29 |
| **Step 4: psychological factors** | $(\vert R^2\vert = .572, p < .001; \Delta R^2 = .192; p < .001)$ | | $(\vert R^2\vert = .575, p < .001; \Delta R^2 = .207; p < .001)$ | | $(\vert R^2\vert = .582, p < .001; \Delta R^2 = .258; p < .001)$ | |
| Depression (PHQ-9) | **.469** | **<.001** | **.385** | **<.001** | **.459** | **<.001** |
| Anxiety (GAD-7) | .066 | .24 | **.171** | **.004** | **.166** | **.003** |
| Adverse Childhood Experience (ACE) | .015 | .69 | .067 | .07 | .049 | .19 |

Note. Coefficients from the final regression model are reported in each step. Due to missing values, the following sample sizes result in the individual groups: n CKD group = 359, n CHD group = 414, n healthy control group = 363. Bold types stand for significant values.
CKD, chronic kidney disease; CHD, coronary heart disease; eGFR, estimated glomerular filtration rate; BMI, Body-Mass-Index; PHQ-9, Patient-Health-Questionnaire-9; GAD-7, Generalized-Anxiety-Disorder-7; ACE, Adverse Childhood Experiences; $R^2$ , coefficient of determination $R^2$ ; $\Delta R^2$ , adjusted $R^2$ ; p, p-value;

**Table 3:**
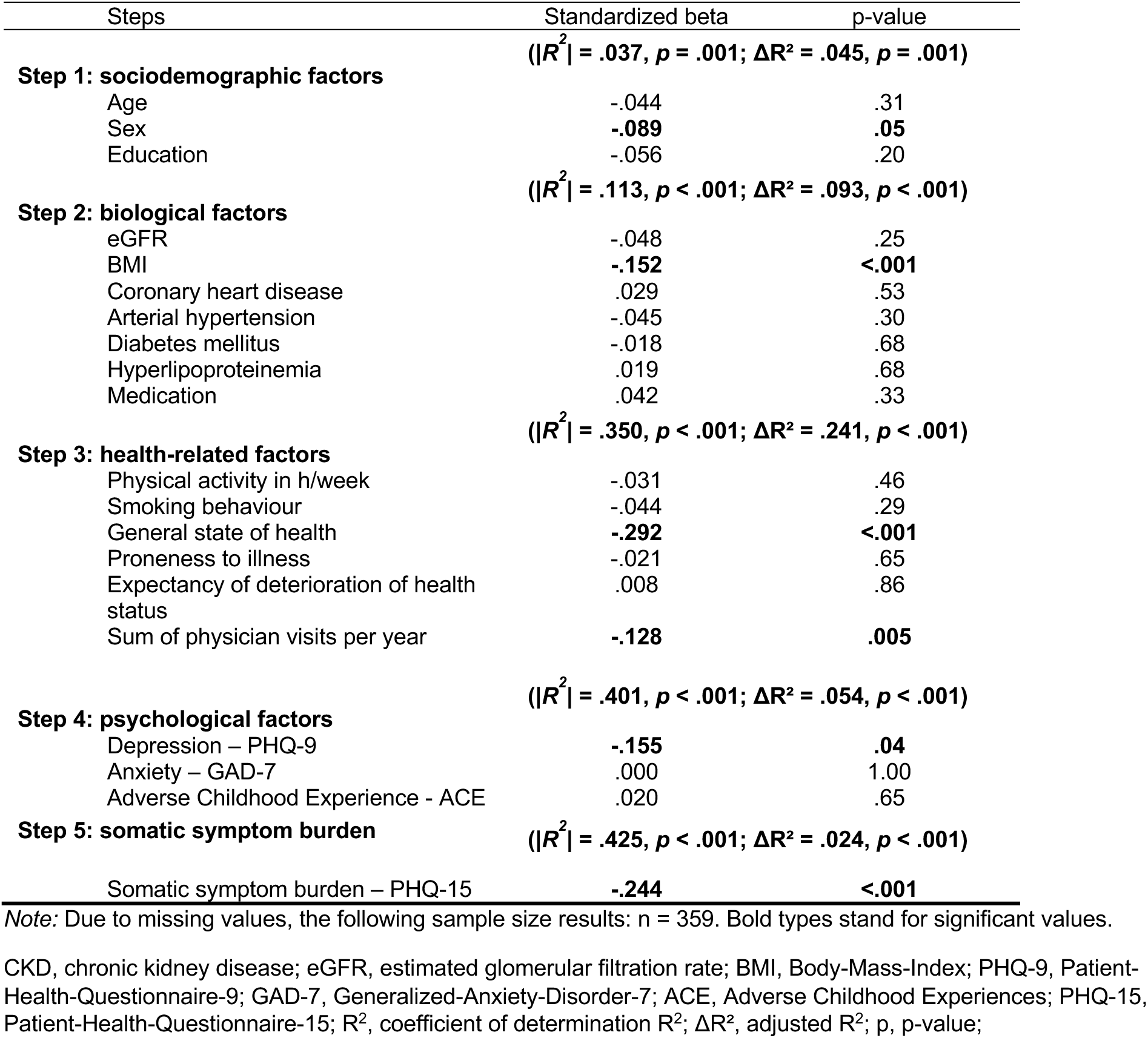
Correlates of quality of life in individuals with CKD using a stepwise multiple linear regression model.

| Steps | Standardized beta | p-value |
| --- | --- | --- |
| | $(\vert R^2\vert = .037, p = .001; \Delta R^2 = .045, p = .001)$ | |
| **Step 1: sociodemographic factors** |  |  |
| Age | -.044 | .31 |
| Sex | **-.089** | **.05** |
| Education | -.056 | .20 |
| | $(\vert R^2\vert = .113, p < .001; \Delta R^2 = .093, p < .001)$ | |
| **Step 2: biological factors** |  |  |
| eGFR | -.048 | .25 |
| BMI | **-.152** | **<.001** |
| Coronary heart disease | .029 | .53 |
| Arterial hypertension | -.045 | .30 |
| Diabetes mellitus | -.018 | .68 |
| Hyperlipoproteinemia | .019 | .68 |
| Medication | .042 | .33 |
| | $(\vert R^2\vert = .350, p < .001; \Delta R^2 = .241, p < .001)$ | |
| **Step 3: health-related factors** |  |  |
| Physical activity in h/week | -.031 | .46 |
| Smoking behaviour | -.044 | .29 |
| General state of health | **-.292** | **<.001** |
| Proneness to illness | -.021 | .65 |
| Expectancy of deterioration of health status | .008 | .86 |
| Sum of physician visits per year | **-.128** | **.005** |
| | $(\vert R^2\vert = .401, p < .001; \Delta R^2 = .054, p < .001)$ | |
| **Step 4: psychological factors** |  |  |
| Depression – PHQ-9 | **-.155** | **.04** |
| Anxiety – GAD-7 | .000 | 1.00 |
| Adverse Childhood Experience - ACE | .020 | .65 |
| | $(\vert R^2\vert = .425, p < .001; \Delta R^2 = .024, p < .001)$ | |
| **Step 5: somatic symptom burden** |  |  |
| Somatic symptom burden – PHQ-15 | **-.244** | **<.001** |

Note: Due to missing values, the following sample size results: n = 359. Bold types stand for significant values.
CKD, chronic kidney disease; eGFR, estimated glomerular filtration rate; BMI, Body-Mass-Index; PHQ-9, Patient-Health-Questionnaire-9; GAD-7, Generalized-Anxiety-Disorder-7; ACE, Adverse Childhood Experiences; PHQ-15, Patient-Health-Questionnaire-15; $R^2$ , coefficient of determination $R^2$ ; $\Delta R^2$ , adjusted $R^2$ ; p, p-value;

### Biopsychosocial correlates of quality of life

Higher somatic symptom burden correlated with lower QoL in the CKD group (*r* = -.538, *p* < .001). Additionally, all except one single item of the PHQ-15 correlated significantly and negatively with QoL (S2). In the multivariable regression model, male sex (β = -.089, *p* = .05), lower BMI (β = -.152, *p* < .001), better general state of health (β = -.292, *p* < .001), fewer physician visits per year (β = -.128, *p* = .005), as well as lower somatic symptom burden (β = -.244, *p* < .001) were associated with higher QoL. The final regression model explained 42.5% of the variance in QoL.

### Comparison of biopsychosocial correlates of somatic symptom burden across CKD, CHD and healthy controls

Frequencies of individual PHQ-15 symptoms were compared between groups. Individuals with CKD and healthy controls experienced less symptoms than the CHD group. Symptom frequency differed for the following symptoms, among others: *stomach* or *chest pain*, *shortness of breath* and *lack of energy* (S3). Odds ratios calculated with logistic regression models showed that *stomach complaints* as well as *cardiac symptoms* were less frequent in the CKD group compared to the CHD group.

Comparing biopsychosocial correlates of somatic symptom burden in individuals with CKD to matched healthy and somatic controls with CHD, analogous stepwise multiple linear regression models were calculated in the two control groups (table 2). Common correlates of somatic symptom burden across all three groups were higher depressive symptoms (β = .385 to .469, *p*s < .001), lower general health state (β = .117 to .201, *p*s = .006 to < .001), and partly proneness to illness (β = -.077 to -.076, *p*s = .05 to .04). Female sex was associated with higher symptom burden only in the CKD and CHD group (β = .143 and .129, *p*s < .001). Higher anxiety levels were associated with higher symptom burden in the CHD (β = .171, *p*s = .004) and the healthy control group (β = .166, *ps* .003). Unique correlates of somatic symptom burden in the CHD group were age (β = .150, *p* < .001), and sum of physician visits per year (β = .088, *p* = .01). In the healthy control group, higher eGFR (β = .080, *p* = .03) and expectancy of deterioration of health status (β = -.120, *p* = .001) were identified as unique associates of somatic symptom burden.

### Comparison of sociodemographic and clinical variables across CKD stages

GFR stages showed a significant positive correlation with somatic symptom burden across stages 2 to 5 (*r* = .137, *p* = .002. Tables S4 and S5 provide an overview of social demographic and clinical variables as well as symptom frequencies across CKD stages 3a-5. Due to a little number of subjects in stage 4 and 5, stages were summarised as follows: stage 3a, stage 3b to 5. When comparing psychological assessments, the groups differed particularly in terms of somatic symptom burden and QoL. Individuals in stage 3a reported a lower symptom burden (*mean* = 5.37, *SD* = 4.19 vs. *mean* = 6.78, *SD* = 4.20, *p* = .003) and a higher quality of life (*mean* = 0.88, *SD* = 0.16 vs. *mean* = 0.84, *SD* = 0.15, *p* = .01) than individuals in stages 3b to 5.

## Discussion

This study examined biopsychosocial correlates of somatic symptom burden in individuals with CKD. Explaining 57.2% of its variance, significant correlates included female sex, the comorbid existence of coronary heart disease, subjectively perceived lower general health status and higher depressive symptoms.

This is consistent with existing research, identifying female sex as risk factor for PSS across diseases^42,43^, including CKD.^7^ In line with a recent study, the presence of coronary heart disease seemed to explain more of the symptom burden than kidney function.^7^ Possible reasons might be that cardiovascular complications are closely linked to CKD and are considered one of the main causes of death in individuals with CKD.^44^ As in this study, patients with CHD generally report a high symptom burden.^10^

Although associated bivariately, the eGFR was not associated with higher somatic symptom burden in the multivariable regression model, a finding supported by recent research showing that eGFR was only associated with individual symptoms but not overall symptom burden.^7^ Our data further confirmed that somatic symptom burden has a strong impact on QoL ^45,46^ by finding a negative correlation between QoL and high symptom burden (see S1). QoL has been identified as associative factor of symptom burden in CKD before, especially in end-stage kidney disease.^8,15,47^ Even if CKD is considered asymptomatic until advanced stages in past studies, patients in this study report various symptoms such as *pain in arms, legs or joints with a frequency* of up to 64.3%.^7,8^

Depressive symptoms, anxiety, and adverse childhood experiences were univariately correlated with somatic symptom burden and QoL. However, depressive symptoms emerged as the only significant psychological correlate in the regression model, even if all factors contributed to symptom persistence in past studies.^1,2,15–17^.

We further aimed to compare correlates of somatic symptom burden in CKD with CHD and healthy controls. Common correlates across all three groups included higher depressive symptoms and lower general health status. Surprisingly, anxiety did not seem to play a role in somatic symptom burden in CKD, but it appeared to be an important correlate for higher symptom burden in both of the control groups. One possible explanation could be that anxiety is more common in patients with CHD when comparing to the general population, and anxiety is a symptom genuinely associated with cardiac conditions.^48^ Considering the general symptom burden and single symptoms of the PHQ-15, the CHD group showed the highest values in our sample. This could be due to the aforementioned general symptom burden of this patient group.^10^

Similar risk and protective factors for somatic symptom distress in the three groups were found, while some variables were specific correlates in only one group. Putting this into perspective in the biopsychosocial model of PSS, it is in line with broad evidence both in CKD and in other chronic conditions, as there are disease-overarching (e.g. female sex, depression) as well as disease-specific correlates (e.g. kidney function) with somatic symptom burden.^1,2,15–17^ The relevance of depression and anxiety as risk factors for PSS have broadly been shown in previous research^2,13,14^, and were confirmed in our analysis.

Thus, the results support a disease-overarching concept of PSS, stressing the important observation that - across all fields of medicine - symptoms can be the primary burden for patients.^1^ The complete proposed biopsychosocial model cannot be tested within the limited set of variables assessed in the HCHS. Other relevant psychosocial factors not assessed here include patients’ symptom and treatment expectations and experiences, cognitive-perceptual and emotional mechanisms such as catastrophizing, somatosensory amplification and emotion regulations deficits.^1,4^ Novel predictive processing models interpret symptoms as imprecise representations of bodily states, often more shaped by priors (e.g. previous symptom experience) than by actual somatosensory input.^2^ Crucially, these overarching mechanisms provide a convincing basis for a mechanism-driven, transdiagnostic, symptom-centered treatment approach. They should therefore be systematically integrated into the treatment and into a comprehensive biopsychosocial model of CKD.

While the HCHS provides a large representative data pool analysing biopsychosocial correlates of somatic symptom burden and allows for a comparison of CKD with another symptomatic disease (CHD) and a healthy control group, several limitations need to be considered: The cross-sectional design of the analysis does not allow for causal inferences. Only general symptoms were assessed, while CKD-specific symptom questionnaires such as the *Kidney Disease Quality of Life Instrument* (KDQOL)^49^ or the *Chronic Kidney Disease Symptom Burden Index* (CKD-SBI)^50^ would have provided more information on CKD-specific symptom burden. Unfortunately, CKD-specific questionnaires were not assessed as the HCHS study has agreed on a generic core set of questionnaires. Due to a limited number of correlates, other health-related and psychosocial factors such as illness perceptions or negative affectivity were not analysed in this study, which were identified as predictors of non-adherence, depression, morbidity and mortality in patients with CKD in former research.^19,23–25^ Due to several single missing values in the sociodemographic data and scales, our sample for the regression analysis was reduced, limiting result validity. Another limitation was the small number of individuals with advanced CKD.

In conclusion, our study shows that symptom burden is only marginally related to kidney function in individuals with CKD, while other biopsychosocial factors such as comorbid cardiac disease and depressive symptoms play a more important role. More research on influencing factors and their underlying aetiology of PSS in CKD is necessary. To gain deeper insights into all aspects of symptom burden in CKD, it is necessary to consider a biopsychosocial model of symptom burden in CKD establishing interdisciplinary treatment approaches for PSS in CKD. As recommended in the KDIGO guidelines^29^ a more interdisciplinary care in the treatment of CKD in future is postulated. This means that in addition to nephrologists or other specialists, nutritionists and psychologists should be more involved in the treatment improving symptom assessment and management.^27^ The SOMA.CK study is investigating biopsychosocial predictors of symptom burden in non-dialysis CKD, aiming to identify modifiable factors and to form future interventions to reduce symptom burden, emphasizing the importance of a biopsychosocial model to better understand and influence treatment outcomes.^4^

## Data Availability

Data will be shared upon reasonable request to the corresponding author. Data-sharing is subject to the approval of the steering committee of the Hamburg City Health Study.

## Acknowledgements

We are grateful for the HCHS team for their support with the data. We thank the involved student research assistants from the Medical School Hamburg. During the preparation of this work the authors used DEEPL in the writing process for better readability. After using this tool, the authors reviewed and edited the content as needed and take full responsibility for the content of the publication.

## CRediT authorship contribution statement

**Birte Jessen:** conceptualization, methodology, formal analysis, writing (original draft & review and editing). **Meike Shedden-Mora:** conceptualization, methodology, formal analysis, writing (original draft & review and editing), supervision. **Tobias B. Huber:** conceptualization, writing (review & editing), methodology, supervision. **Christian Schmidt-Lauber:** writing (review & editing), methodology, supervision. **Bernd Löwe:** conceptualization, writing (review & editing), methodology. **Raphael Twerenbold:** writing (review & editing). **Martin Härter:** writing (review & editing). **Volker Harth:** writing (review & editing). **Hanno Hoven:** writing (review & editing).

## Supplemental material

Supplemental material is available online at …

## Supplements

**Table S1:**
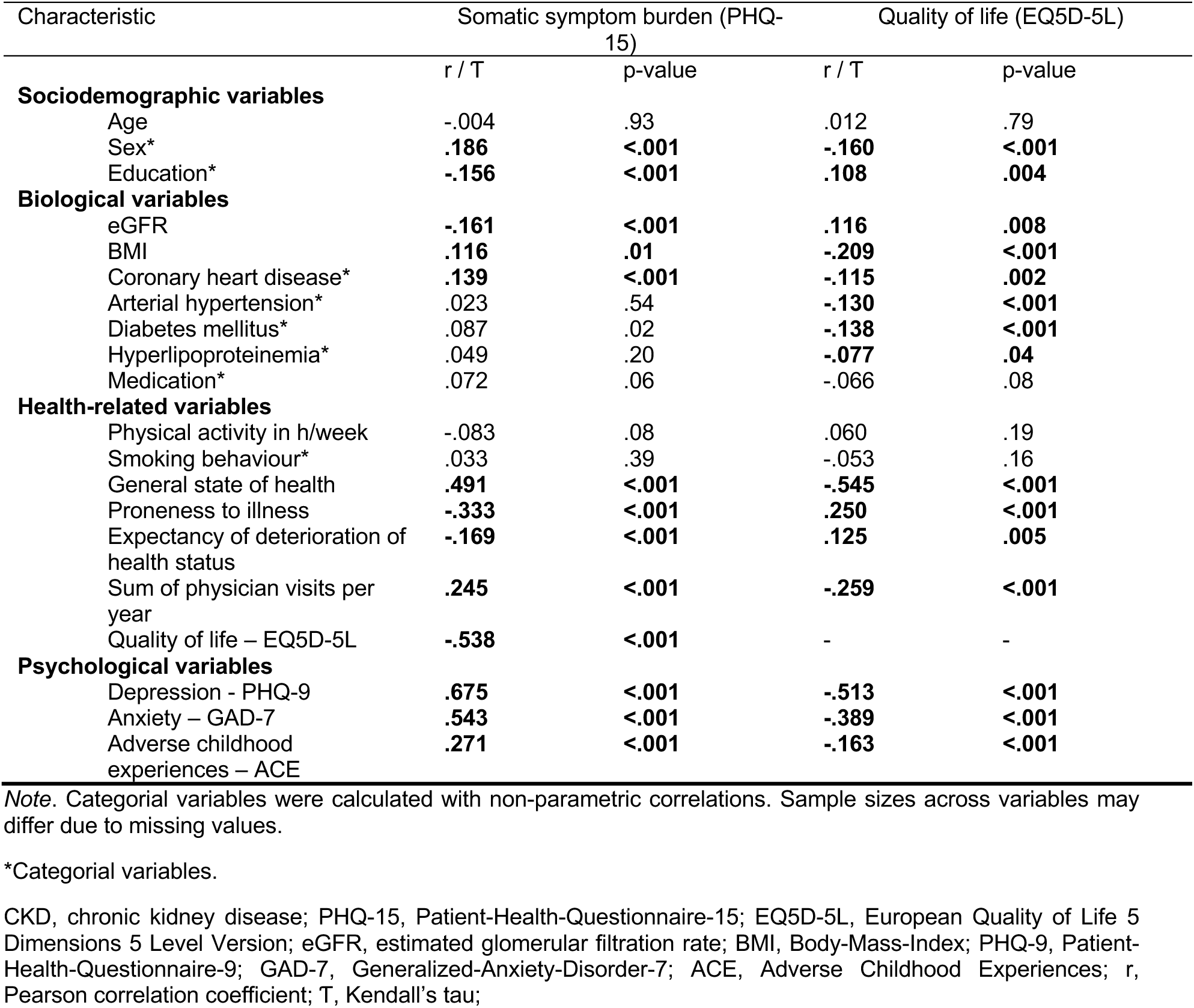
Correlations with somatic symptom burden and quality of life in individuals with CKD.

| Characteristic | Somatic symptom burden (PHQ-15) |  | Quality of life (EQ5D-5L) |  |
| --- | --- | --- | --- | --- |
|  | r / T | p-value | r / T | p-value |
| **Sociodemographic variables** |  |  |  |  |
| Age | -.004 | .93 | .012 | .79 |
| Sex* | **.186** | **<.001** | **-.160** | **<.001** |
| Education* | **-.156** | **<.001** | **.108** | **.004** |
| **Biological variables** |  |  |  |  |
| eGFR | **-.161** | **<.001** | **.116** | **.008** |
| BMI | **.116** | **.01** | **-.209** | **<.001** |
| Coronary heart disease* | **.139** | **<.001** | **-.115** | **.002** |
| Arterial hypertension* | .023 | .54 | **-.130** | **<.001** |
| Diabetes mellitus* | .087 | .02 | **-.138** | **<.001** |
| Hyperlipoproteinemia* | .049 | .20 | **-.077** | **.04** |
| Medication* | .072 | .06 | -.066 | .08 |
| **Health-related variables** |  |  |  |  |
| Physical activity in h/week | -.083 | .08 | .060 | .19 |
| Smoking behaviour* | .033 | .39 | -.053 | .16 |
| General state of health | **.491** | **<.001** | **-.545** | **<.001** |
| Proneness to illness | **-.333** | **<.001** | **.250** | **<.001** |
| Expectancy of deterioration of health status | **-.169** | **<.001** | **.125** | **.005** |
| Sum of physician visits per year | **.245** | **<.001** | **-.259** | **<.001** |
| Quality of life – EQ5D-5L | **-.538** | **<.001** | - | - |
| **Psychological variables** |  |  |  |  |
| Depression - PHQ-9 | **.675** | **<.001** | **-.513** | **<.001** |
| Anxiety – GAD-7 | **.543** | **<.001** | **-.389** | **<.001** |
| Adverse childhood experiences – ACE | **.271** | **<.001** | **-.163** | **<.001** |

*Note.* Categorical variables were calculated with non-parametric correlations. Sample sizes across variables may differ due to missing values.
\*Categorical variables.
CKD, chronic kidney disease; PHQ-15, Patient-Health-Questionnaire-15; EQ5D-5L, European Quality of Life 5 Dimensions 5 Level Version; eGFR, estimated glomerular filtration rate; BMI, Body-Mass-Index; PHQ-9, Patient-Health-Questionnaire-9; GAD-7, Generalized-Anxiety-Disorder-7; ACE, Adverse Childhood Experiences; r, Pearson correlation coefficient; T, Kendall's tau;

**Table S2:** Correlations with quality of life and single somatic symptoms in individuals with CKD.

| Characteristic | Quality of life (EQ5D-5L) |  |
| --- | --- | --- |
| PHQ-15 variables | r | p-value |
| Stomach pain | **-.145** | **.001** |
| Back pain | **-.455** | **<.001** |
| Pain in your arms, legs, or joints (knee, hips, etc.) | **-.546** | **<.001** |
| Menstrual cramps or other problems with your periods (women only) | -.080 | .07 |
| Pain or problems during sexual intercourse | **-.136** | **.002** |
| Headaches | **-.263** | **<.001** |
| Chest pain | **-.265** | **<.001** |
| Dizziness | **-.275** | **<.001** |
| Fainting spells | **-.176** | **<.001** |
| Feeling your heart pound or race | **-.165** | **<.001** |
| Shortness of breath | **-.352** | **<.001** |
| Constipation, loose bowels, or diarrhoea | **-.179** | **<.001** |
| Nausea, gas, or indigestion | **-.212** | **<.001** |
| Feeling tired or having low energy | **-.420** | **<.001** |
| Trouble sleeping | **-.266** | **<.001** |

*Note:* Sample sizes across variables may differ due to missing values.
CKD, chronic kidney disease; PHQ-15, Patient-Health-Questionnaire-15; EQ5D-5L, European Quality of Life 5 Dimensions 5 Level Version; r, Pearson correlation coefficient;

**Table S3:** Symptom amount measured with PHQ-15 – comparison of groups.

| Characteristic | cohort of 10.000 | CKD group<sup>a</sup> | CHD group<sup>b</sup> | healthy control group<sup>c</sup> | comparison | CKD vs. CHD OR, 95% CI | CKD vs. HC OR, 95% CI |
| --- | --- | --- | --- | --- | --- | --- | --- |
| n | 10.000 | 582 | 618 | 582 |  |  |  |
| **PHQ-15 variables** |  |  |  |  |  |  |  |
| Stomach Pain n (%) | 1508 (15.1%) | 73 (12.5%) | 112 (18.1%) | 98 (16.8%) | $\chi^2 = 7.58, p = .02$<br>**(a < b, c)** | 0.65 [.471, .892]<br>**(a < b)** | 0.71 [.510, .983]<br>**(a < c)** |
| Back Pain n (%) | 4832 (48.3%) | 304 (52.2%) | 323 (52.3%) | 302 (51.9%) | $\chi^2 = 0.02, p = .99$ | 1.00 [.796, 1.253] | 1.01 [.806, 1.276] |
| Pain in your arms, legs, or joints (knee, hips, etc.) n (%) | 5328 (53.3%) | 360 (61.9%) | 372 (60.2%) | 336 (57.7%) | $\chi^2 = 2.09, p = .35$ | 1.07 [.850, 1.353] | 1.19 [.939, 1.501] |
| Menstrual cramps or other problems with your periods (women only) n (%) | 283 (2.8%) | 4 (0.7%) | 2 (0.3%) | 6 (1.0%) | $\chi^2 = 2.24, p = .33$ | 2.13 [.389, 11.68] | 0.66 [.186, 2.367] |
| Pain or problems during sexual intercourse n (%) | 1567 (15.7%) | 127 (21.8%) | 149 (24.1%) | 107 (18.4%) | $\chi^2 = 5.88, p = .05$<br>**(b > c)** | 0.88 [.671, 1.150] | 1.24 [.929, 1.652] |
| Headaches n (%) | 2733 (27.3%) | 118 (20.3%) | 141 (22.8%) | 124 (21.3%) | $\chi^2 = 1.16, p = .56$ | 0.86 [.653, 1.134] | 0.94 [.708, 1.247] |
| Chest pain n (%) | 1064 (10.6%) | 67 (11.5%) | 145 (23.5%) | 55 (9.5%) | $\chi^2 = 54.38, p < .001$<br>**(a < b; b > c)** | 0.43 [.310, .582]<br>**(a < b)** | 1.25 [.855, 1.817] |
| Dizziness n (%) | 2079 (20.8%) | 155 (26.6%) | 179 (29.0%) | 146 (25.1%) | $\chi^2 = 2.33, p = .31$ | 0.89 [.691, 1.147] | 1.08 [.834, 1.409] |
| Fainting spells n (%) | 128 (1.3%) | 9 (1.5%) | 11 (1.8%) | 14 (2.4%) | $\chi^2 = 1.23, p = .54$ | 0.87 [.357, 2.107] | 0.64 [.274, 1.484] |
| Feeling your heart pound or race n (%) | 1701 (17%) | 114 (19.6%) | 183 (29.6%) | 96 (16.5%) | $\chi^2 = 33.06, p < .001$<br>**(a < b; b > c)** | 0.58 [.443, .757]<br>**(a < b)** | 1.23 [.914, 1.664] |
| Shortness of breath n (%) | 2518 (25.2%) | 210 (36.1%) | 251 (40.6%) | 157 (27.0%) | $\chi^2 = 25.36, p < .001$<br>**(a, b > c)** | 0.83 [.654, 1.042] | 1.53 [1.191, 1.961]<br>**(a > c)** |
| Constipation, loose bowels, or diarrhoea n (%) | 2760 (27.6%) | 154 (26.5%) | 205 (33.2%) | 163 (28.0%) | $\chi^2 = 7.21, p = .03$<br>**(a < b)** | 0.73 [.565, .930]<br>**(a < b)** | 0.93 [.714, 1.197] |
| Nausea, gas, or indigestion n (%) | 3103 (31%) | 158 (27.1%) | 219 (35.4%) | 177 (30.4%) | $\chi^2 = 9.80, p = .007$<br>**(a < b)** | 0.68 [.531, .868]<br>**(a < b)** | 0.85 [.661, 1.099] |
| Feeling tired or having low energy n (%) | 4601 (46%) | 258 (44.3%) | 283 (45.8%) | 233 (40.0%) | $\chi^2 = 4.33, p = .12$<br>**(b > c)** | 0.94 [.751, 1.183] | 1.19 [.945, 1.506] |
| Trouble sleeping n (%) | 4800 (48%) | 270 (46.4%) | 307 (49.7%) | 264 (45.4%) | $\chi^2 = 2.46, p = .29$ | 0.88 [.699, 1.100] | 1.04 [.828, 1.313] |
| Symptom amount (mean ± SD) | 4.27 ± 3.02 | 4.69 ± 3.11 | 5.19 ± 3.50 | 4.32 ± 3.08 | $F = 9.99, p < .001$<br>**(a < b; b > c)** | - | - |

Note: Sample sizes across variables may differ due to missing values. Bold types stand for significant values.
PHQ-15, Patient-Health-Questionnaire-15; CKD, chronic kidney disease; CHD, coronary heart disease; OR, Odds ratio; CI, confidence interval; n, number; $\chi^2$ , chi-square coefficient; p, p-value; F, F-value;

**Table S4:**
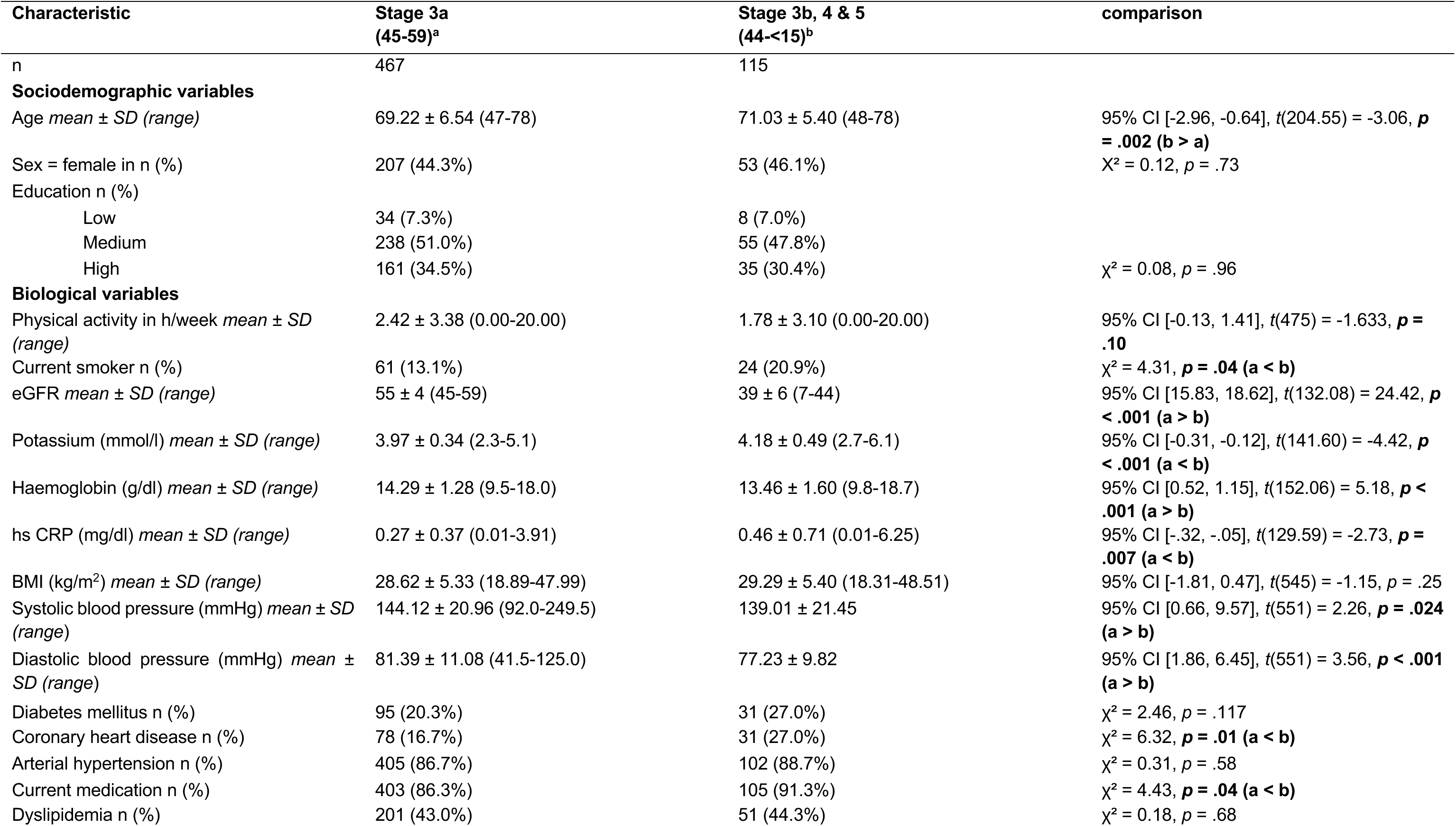
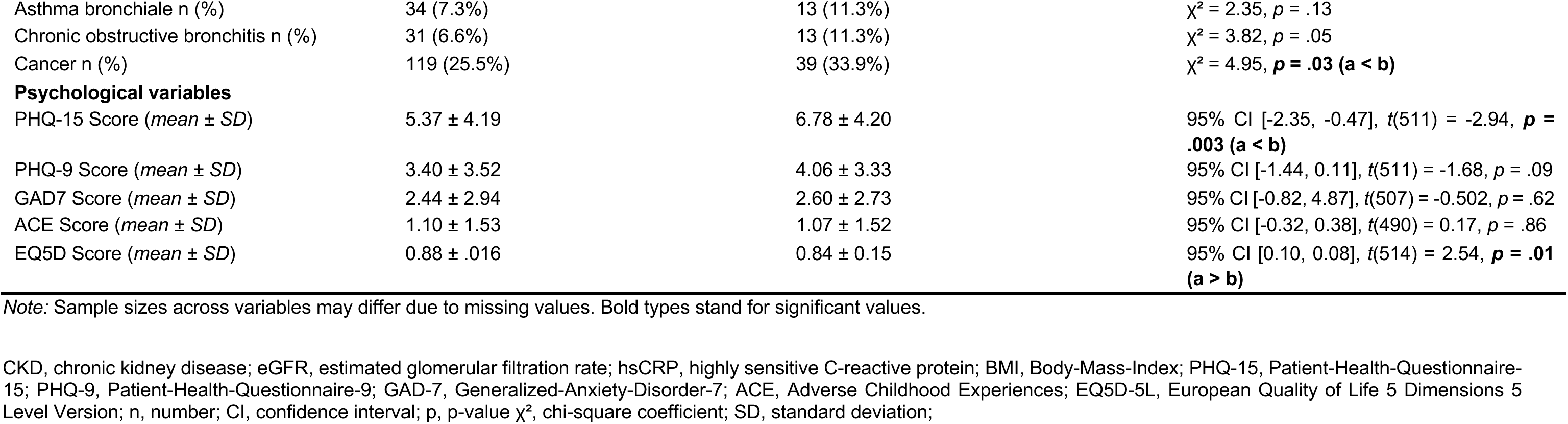
Comparison of social demographic and clinical variables across CKD stages.

| Characteristic | Stage 3a<br>(45-59)<sup>a</sup> | Stage 3b, 4 & 5<br>(44-<15)<sup>b</sup> | comparison |
| --- | --- | --- | --- |
| n | 467 | 115 |  |
| **Sociodemographic variables** |  |  |  |
| Age mean ± SD (range) | 69.22 ± 6.54 (47-78) | 71.03 ± 5.40 (48-78) | 95% CI [-2.96, -0.64], $t(204.55) = -3.06$, **$p = .002$ (b > a)** |
| Sex = female in n (%) | 207 (44.3%) | 53 (46.1%) | $\chi^2 = 0.12$, $p = .73$ |
| Education n (%) |  |  |  |
| Low | 34 (7.3%) | 8 (7.0%) |  |
| Medium | 238 (51.0%) | 55 (47.8%) |  |
| High | 161 (34.5%) | 35 (30.4%) | $\chi^2 = 0.08$, $p = .96$ |
| **Biological variables** |  |  |  |
| Physical activity in h/week mean ± SD (range) | 2.42 ± 3.38 (0.00-20.00) | 1.78 ± 3.10 (0.00-20.00) | 95% CI [-0.13, 1.41], $t(475) = -1.633$, **$p = .10$** |
| Current smoker n (%) | 61 (13.1%) | 24 (20.9%) | $\chi^2 = 4.31$, **$p = .04$ (a < b)** |
| eGFR mean ± SD (range) | 55 ± 4 (45-59) | 39 ± 6 (7-44) | 95% CI [15.83, 18.62], $t(132.08) = 24.42$, **$p < .001$ (a > b)** |
| Potassium (mmol/l) mean ± SD (range) | 3.97 ± 0.34 (2.3-5.1) | 4.18 ± 0.49 (2.7-6.1) | 95% CI [-0.31, -0.12], $t(141.60) = -4.42$, **$p < .001$ (a < b)** |
| Haemoglobin (g/dl) mean ± SD (range) | 14.29 ± 1.28 (9.5-18.0) | 13.46 ± 1.60 (9.8-18.7) | 95% CI [0.52, 1.15], $t(152.06) = 5.18$, **$p < .001$ (a > b)** |
| hs CRP (mg/dl) mean ± SD (range) | 0.27 ± 0.37 (0.01-3.91) | 0.46 ± 0.71 (0.01-6.25) | 95% CI [-.32, -.05], $t(129.59) = -2.73$, **$p = .007$ (a < b)** |
| BMI (kg/m<sup>2</sup>) mean ± SD (range) | 28.62 ± 5.33 (18.89-47.99) | 29.29 ± 5.40 (18.31-48.51) | 95% CI [-1.81, 0.47], $t(545) = -1.15$, $p = .25$ |
| Systolic blood pressure (mmHg) mean ± SD (range) | 144.12 ± 20.96 (92.0-249.5) | 139.01 ± 21.45 | 95% CI [0.66, 9.57], $t(551) = 2.26$, **$p = .024$ (a > b)** |
| Diastolic blood pressure (mmHg) mean ± SD (range) | 81.39 ± 11.08 (41.5-125.0) | 77.23 ± 9.82 | 95% CI [1.86, 6.45], $t(551) = 3.56$, **$p < .001$ (a > b)** |
| Diabetes mellitus n (%) | 95 (20.3%) | 31 (27.0%) | $\chi^2 = 2.46$, $p = .117$ |
| Coronary heart disease n (%) | 78 (16.7%) | 31 (27.0%) | $\chi^2 = 6.32$, **$p = .01$ (a < b)** |
| Arterial hypertension n (%) | 405 (86.7%) | 102 (88.7%) | $\chi^2 = 0.31$, $p = .58$ |
| Current medication n (%) | 403 (86.3%) | 105 (91.3%) | $\chi^2 = 4.43$, **$p = .04$ (a < b)** |
| Dyslipidemia n (%) | 201 (43.0%) | 51 (44.3%) | $\chi^2 = 0.18$, $p = .68$ |
| Asthma bronchiale n (%) | 34 (7.3%) | 13 (11.3%) | $\chi^2 = 2.35, p = .13$ |
| Chronic obstructive bronchitis n (%) | 31 (6.6%) | 13 (11.3%) | $\chi^2 = 3.82, p = .05$ |
| Cancer n (%) | 119 (25.5%) | 39 (33.9%) | $\chi^2 = 4.95, p = .03$ (a < b) |
| **Psychological variables** |  |  |  |
| PHQ-15 Score (mean ± SD) | 5.37 ± 4.19 | 6.78 ± 4.20 | 95% CI [-2.35, -0.47], $t(511) = -2.94, p = .003$ (a < b) |
| PHQ-9 Score (mean ± SD) | 3.40 ± 3.52 | 4.06 ± 3.33 | 95% CI [-1.44, 0.11], $t(511) = -1.68, p = .09$ |
| GAD7 Score (mean ± SD) | 2.44 ± 2.94 | 2.60 ± 2.73 | 95% CI [-0.82, 4.87], $t(507) = -0.502, p = .62$ |
| ACE Score (mean ± SD) | 1.10 ± 1.53 | 1.07 ± 1.52 | 95% CI [-0.32, 0.38], $t(490) = 0.17, p = .86$ |
| EQ5D Score (mean ± SD) | 0.88 ± .016 | 0.84 ± 0.15 | 95% CI [0.10, 0.08], $t(514) = 2.54, p = .01$ (a > b) |

Note: Sample sizes across variables may differ due to missing values. Bold types stand for significant values.
CKD, chronic kidney disease; eGFR, estimated glomerular filtration rate; hsCRP, highly sensitive C-reactive protein; BMI, Body-Mass-Index; PHQ-15, Patient-Health-Questionnaire-15; PHQ-9, Patient-Health-Questionnaire-9; GAD-7, Generalized-Anxiety-Disorder-7; ACE, Adverse Childhood Experiences; EQ5D-5L, European Quality of Life 5 Dimensions 5 Level Version; n, number; CI, confidence interval; p, p-value $\chi^2$ , chi-square coefficient; SD, standard deviation;

**Table S5:** Symptom amount measured with PHQ-15 – comparison of CKD stages.

| Characteristic | Stage 3a<br>(45-59)<sup>a</sup> | Stage 3b, 4 & 5<br>(44-<15)<sup>b</sup> | comparison |
| --- | --- | --- | --- |
| n | 469 | 99 |  |
| **PHQ-15 variables** |  |  |  |
| Stomach Pain n (%) | 57 (12.2%) | 16 (13.9%) | $\chi^2 = .245, p = .62$ |
| Back Pain n (%) | 247 (52.9%) | 57 (49.6%) | $\chi^2 = .409, p = .52$ |
| Pain in your arms, legs, or joints (knee, hips, etc.) n (%) | 286 (61.2%) | 74 (64.3%) | $\chi^2 = .377, p = .54$ |
| Menstrual cramps or other problems with your periods (women only) n (%) | 4 (0.9%) | 0 (0%) | $\chi^2 = .992, p = .32$ |
| Pain or problems during sexual intercourse n (%) | 96 (20.6%) | 31 (27.0%) | $\chi^2 = 2.22, p = .14$ |
| Headaches n (%) | 91 (19.5%) | 27 (23.5%) | $\chi^2 = .910, p = .34$ |
| Chest pain n (%) | 54 (11.6%) | 13 (11.3%) | $\chi^2 = .006, p = .94$ |
| Dizziness n (%) | 120 (25.7%) | 35 (30.4%) | $\chi^2 = 1.06, p = .30$ |
| Fainting spells n (%) | 6 (1.3%) | 3 (2.6%) | $\chi^2 = 1.06, p = .30$ |
| Feeling your heart pound or race n (%) | 83 (17.8%) | 31 (27.0%) | $\chi^2 = 4.94, \mathbf{p = .03 (a < b)}$ |
| Shortness of breath n (%) | 160 (34.3%) | 50 (43.5%) | $\chi^2 = 3.40, p = .07$ |
| Constipation, loose bowels, or diarrhoea n (%) | 115 (24.6%) | 39 (33.9%) | $\chi^2 = 4.09, \mathbf{p = .04 (a < b)}$ |
| Nausea, gas, or indigestion n (%) | 124 (26.6%) | 34 (29.6%) | $\chi^2 = 0.42, p = .52$ |
| Feeling tired or having low energy n (%) | 205 (43.9%) | 53 (46.1%) | $\chi^2 = .18, p = .67$ |
| Trouble sleeping n (%) | 214 (45.8%) | 56 (48.7%) | $\chi^2 = .306, p = .58$ |
| Symptom amount (*mean ± SD*) | 4.49 ± 3.05 | 5.59 ± 3.21 | 95% CI [-1.79, -.040], $t(511) = -3.12, \mathbf{p = .002 (a < b)}$ |

*Note:* Sample sizes across variables may differ due to missing values. Bold types stand for significant values.
CKD, chronic kidney disease; PHQ-15, Patient-Health-Questionnaire-15; n, number; $\chi^2$ , chi-square coefficient; p, p-value; SD, standard deviation; CI, confidence interval;

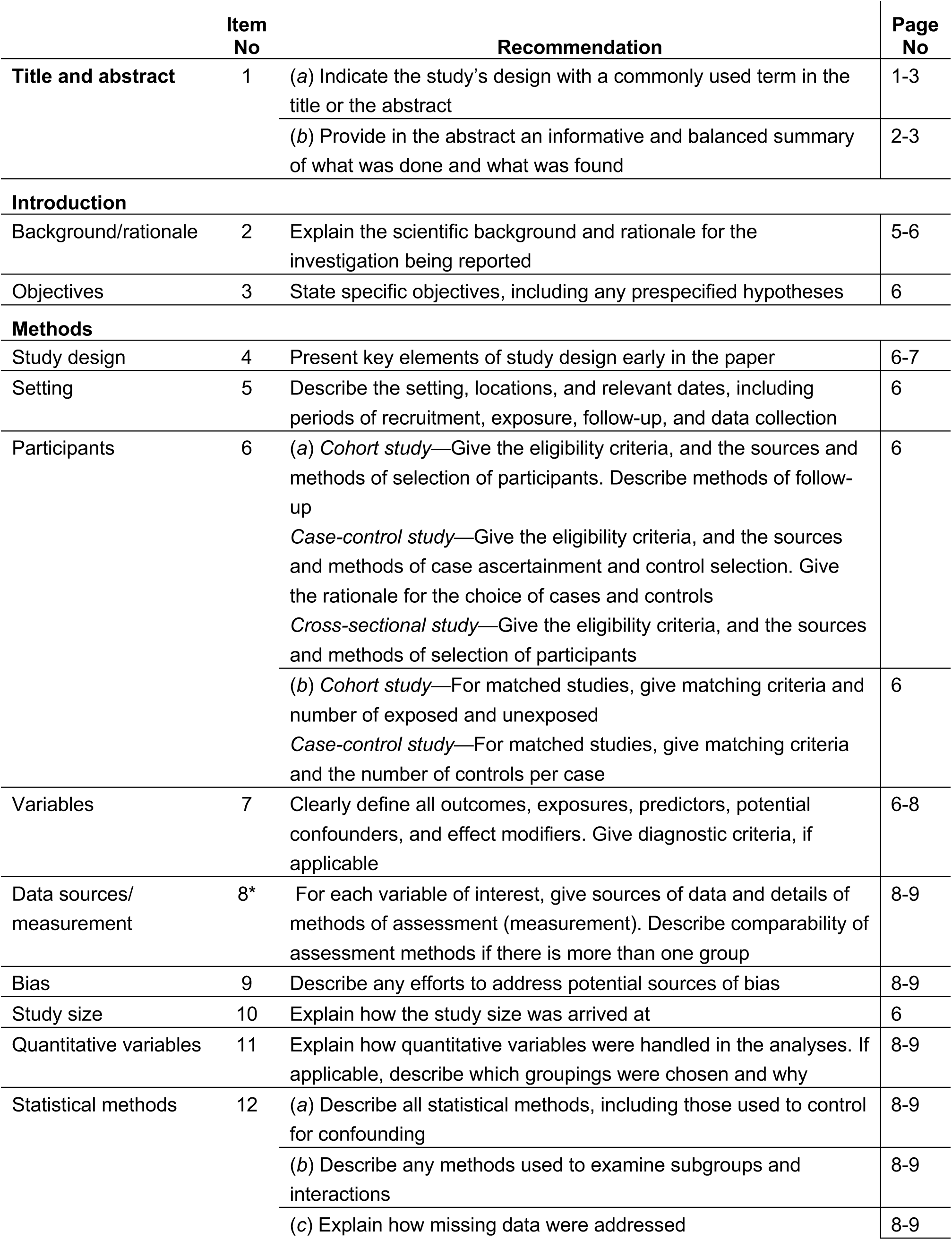
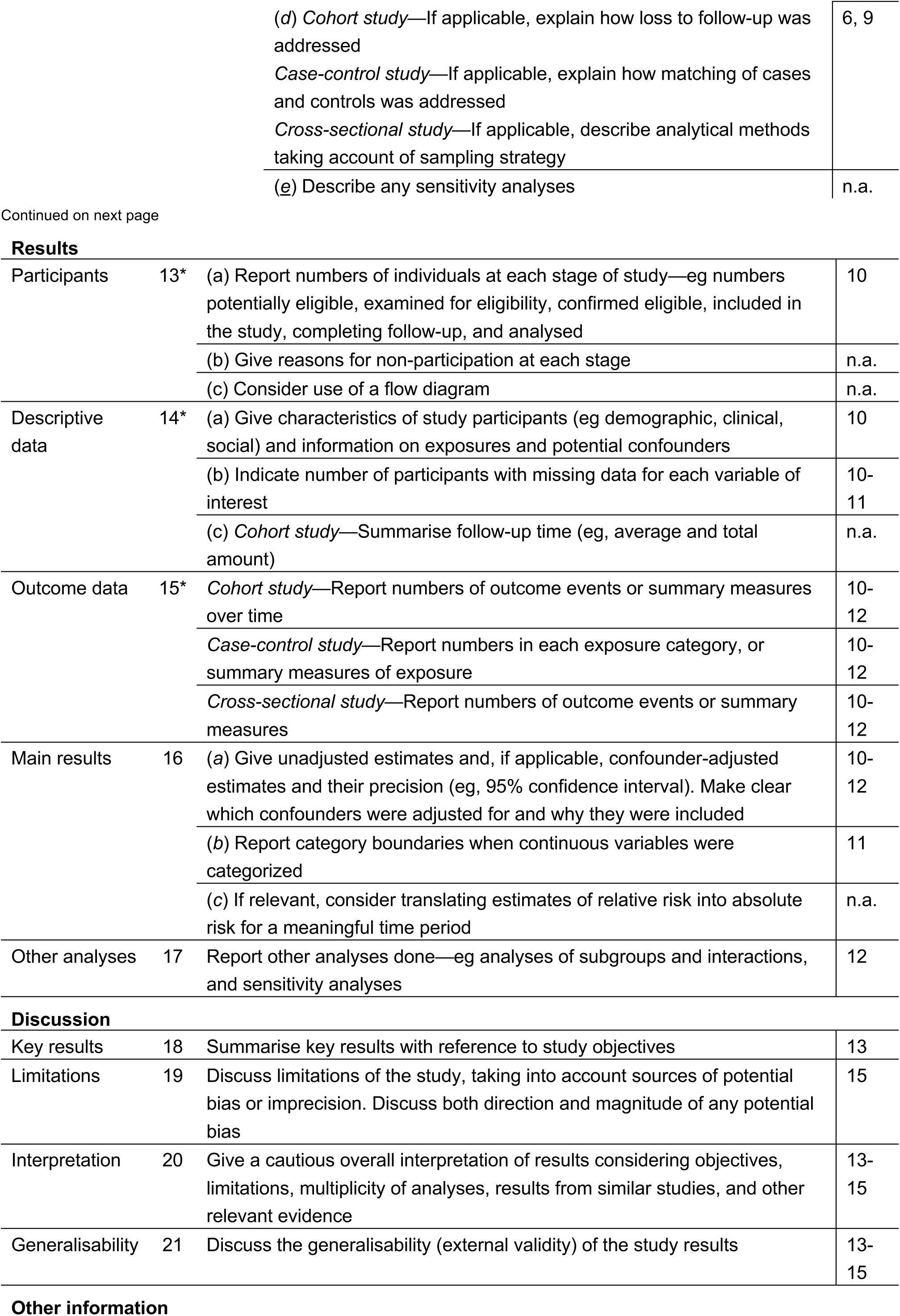
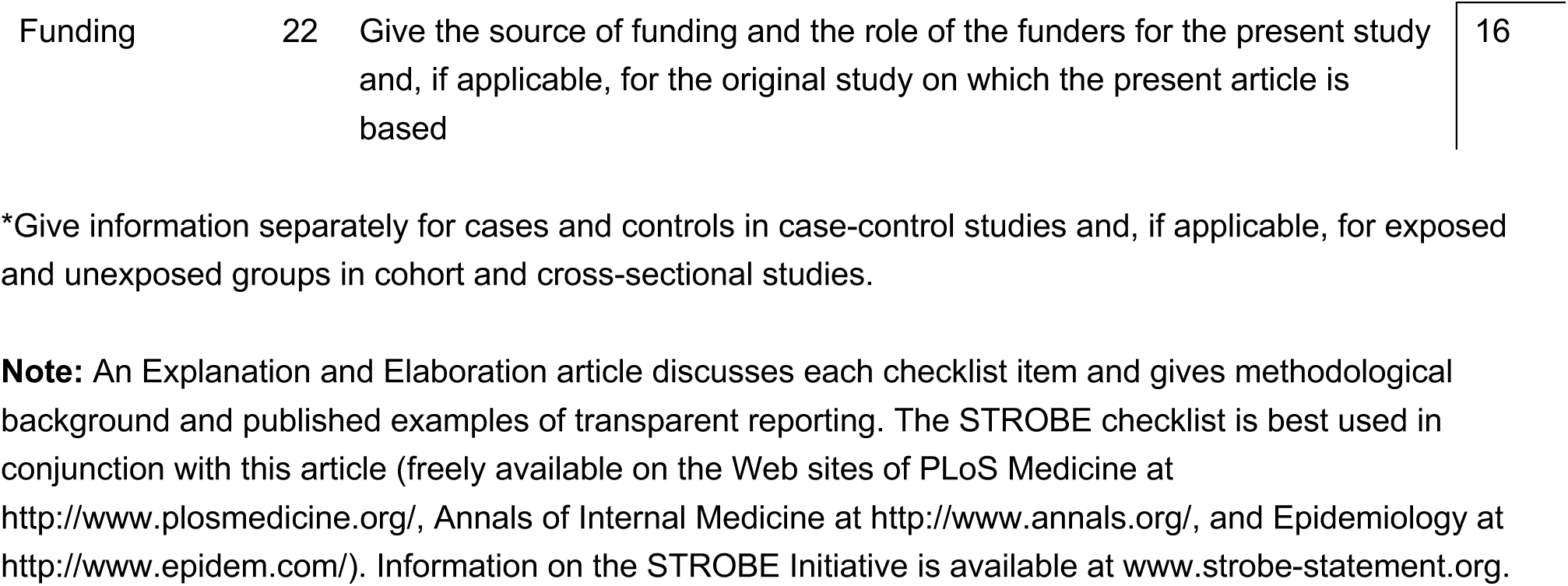
STROBE Statement —checklist of items that should be included in reports of observational studies

|  | Item No | Recommendation | Page No |
| --- | --- | --- | --- |
| Title and abstract | 1 | (a) Indicate the study’s design with a commonly used term in the title or the abstract | 1-3 |
|  |  | (b) Provide in the abstract an informative and balanced summary of what was done and what was found | 2-3 |
| Introduction |  |  |  |
| Background/rationale | 2 | Explain the scientific background and rationale for the investigation being reported | 5-6 |
| Objectives | 3 | State specific objectives, including any prespecified hypotheses | 6 |
| Methods |  |  |  |
| Study design | 4 | Present key elements of study design early in the paper | 6-7 |
| Setting | 5 | Describe the setting, locations, and relevant dates, including periods of recruitment, exposure, follow-up, and data collection | 6 |
| Participants | 6 | (a) Cohort study—Give the eligibility criteria, and the sources and methods of selection of participants. Describe methods of follow-up<br>Case-control study—Give the eligibility criteria, and the sources and methods of case ascertainment and control selection. Give the rationale for the choice of cases and controls<br>Cross-sectional study—Give the eligibility criteria, and the sources and methods of selection of participants | 6 |
|  |  | (b) Cohort study—For matched studies, give matching criteria and number of exposed and unexposed<br>Case-control study—For matched studies, give matching criteria and the number of controls per case | 6 |
| Variables | 7 | Clearly define all outcomes, exposures, predictors, potential confounders, and effect modifiers. Give diagnostic criteria, if applicable | 6-8 |
| Data sources/measurement | 8* | For each variable of interest, give sources of data and details of methods of assessment (measurement). Describe comparability of assessment methods if there is more than one group | 8-9 |
| Bias | 9 | Describe any efforts to address potential sources of bias | 8-9 |
| Study size | 10 | Explain how the study size was arrived at | 6 |
| Quantitative variables | 11 | Explain how quantitative variables were handled in the analyses. If applicable, describe which groupings were chosen and why | 8-9 |
| Statistical methods | 12 | (a) Describe all statistical methods, including those used to control for confounding | 8-9 |
|  |  | (b) Describe any methods used to examine subgroups and interactions | 8-9 |
|  |  | (c) Explain how missing data were addressed | 8-9 |

|  |  |
| --- | --- |
| (d) *Cohort study*—If applicable, explain how loss to follow-up was addressed | 6, 9 |
| *Case-control study*—If applicable, explain how matching of cases and controls was addressed |  |
| *Cross-sectional study*—If applicable, describe analytical methods taking account of sampling strategy |  |
| (e) Describe any sensitivity analyses | n.a. |
| Participants | 13* | (a) Report numbers of individuals at each stage of study—eg numbers potentially eligible, examined for eligibility, confirmed eligible, included in the study, completing follow-up, and analysed | 10 |
|  |  | (b) Give reasons for non-participation at each stage | n.a. |
|  |  | (c) Consider use of a flow diagram | n.a. |
| Descriptive data | 14* | (a) Give characteristics of study participants (eg demographic, clinical, social) and information on exposures and potential confounders | 10 |
|  |  | (b) Indicate number of participants with missing data for each variable of interest | 10-11 |
|  |  | (c) *Cohort study*—Summarise follow-up time (eg, average and total amount) | n.a. |
| Outcome data | 15* | *Cohort study*—Report numbers of outcome events or summary measures over time | 10-12 |
|  |  | *Case-control study*—Report numbers in each exposure category, or summary measures of exposure | 10-12 |
|  |  | *Cross-sectional study*—Report numbers of outcome events or summary measures | 10-12 |
| Main results | 16 | (a) Give unadjusted estimates and, if applicable, confounder-adjusted estimates and their precision (eg, 95% confidence interval). Make clear which confounders were adjusted for and why they were included | 10-12 |
|  |  | (b) Report category boundaries when continuous variables were categorized | 11 |
|  |  | (c) If relevant, consider translating estimates of relative risk into absolute risk for a meaningful time period | n.a. |
| Other analyses | 17 | Report other analyses done—eg analyses of subgroups and interactions, and sensitivity analyses | 12 |
| Key results | 18 | Summarise key results with reference to study objectives | 13 |
| Limitations | 19 | Discuss limitations of the study, taking into account sources of potential bias or imprecision. Discuss both direction and magnitude of any potential bias | 15 |
| Interpretation | 20 | Give a cautious overall interpretation of results considering objectives, limitations, multiplicity of analyses, results from similar studies, and other relevant evidence | 13-15 |
| Generalisability | 21 | Discuss the generalisability (external validity) of the study results | 13-15 |
| Other information |  |  |  |
| Funding | 22 | Give the source of funding and the role of the funders for the present study and, if applicable, for the original study on which the present article is based | 16 |

\*Give information separately for cases and controls in case-control studies and, if applicable, for exposed and unexposed groups in cohort and cross-sectional studies.
**Note:** An Explanation and Elaboration article discusses each checklist item and gives methodological background and published examples of transparent reporting. The STROBE checklist is best used in conjunction with this article (freely available on the Web sites of PLoS Medicine at http://www.plosmedicine.org/, Annals of Internal Medicine at http://www.annals.org/, and Epidemiology at http://www.epidem.com/). Information on the STROBE Initiative is available at www.strobe-statement.org.

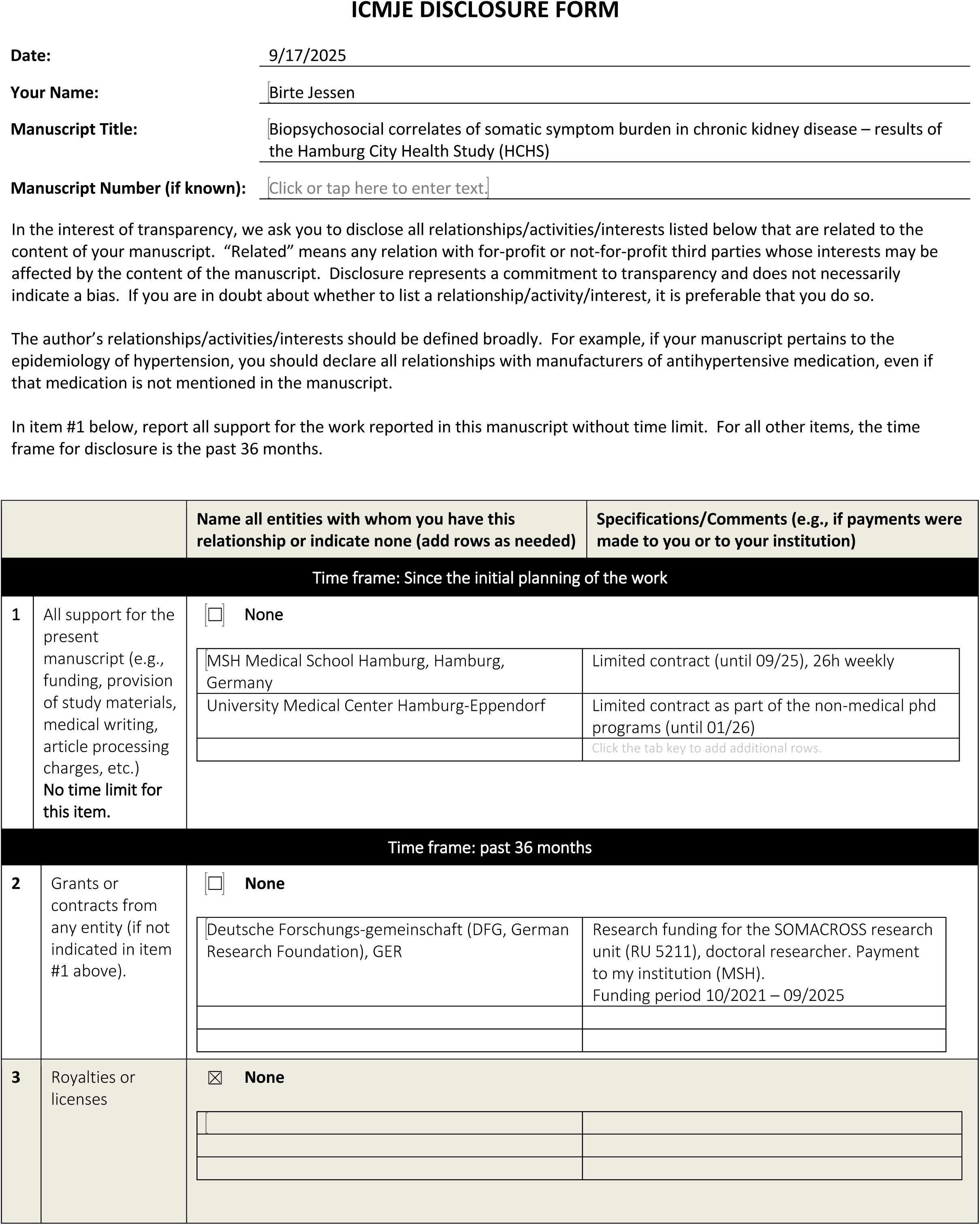
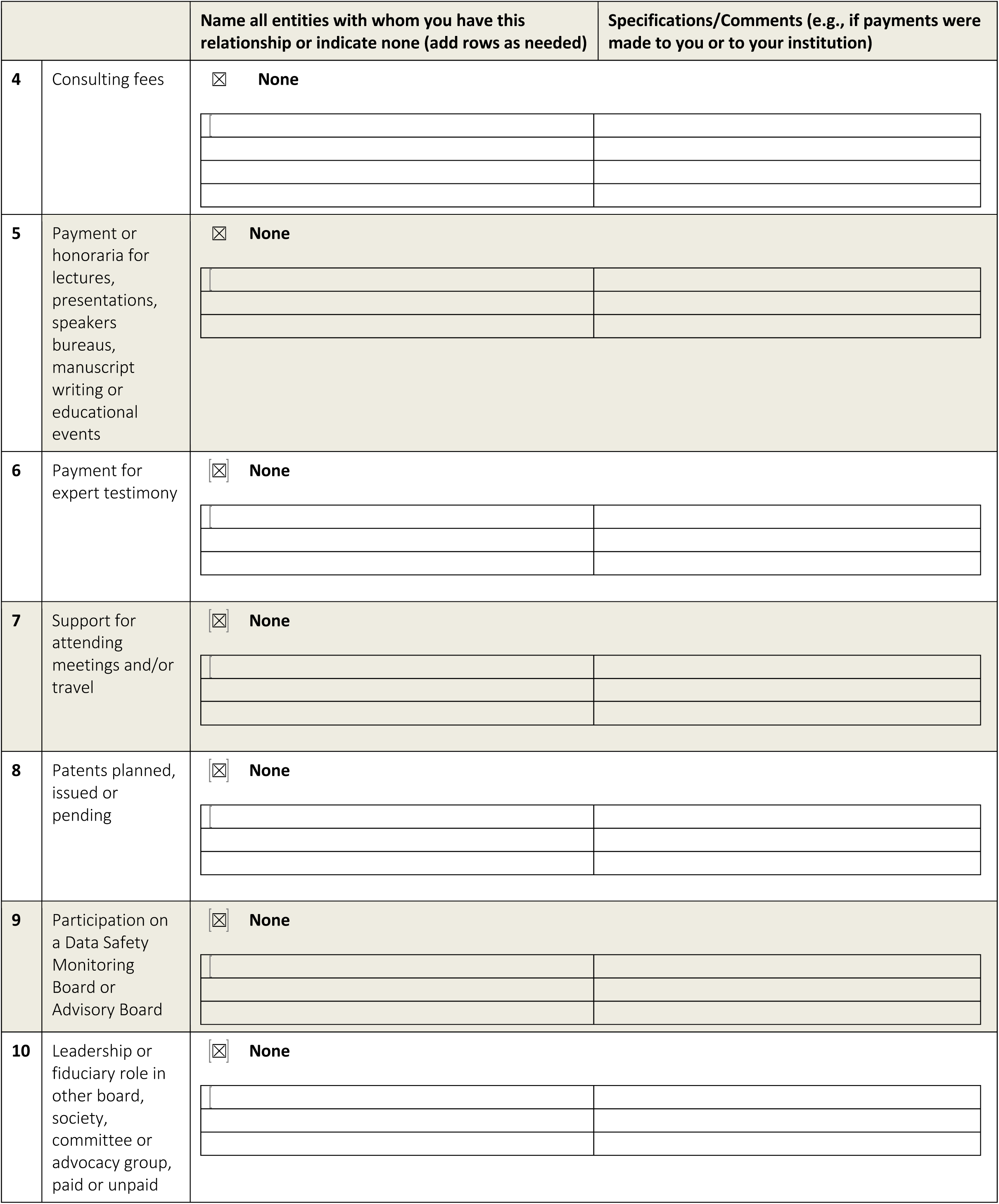
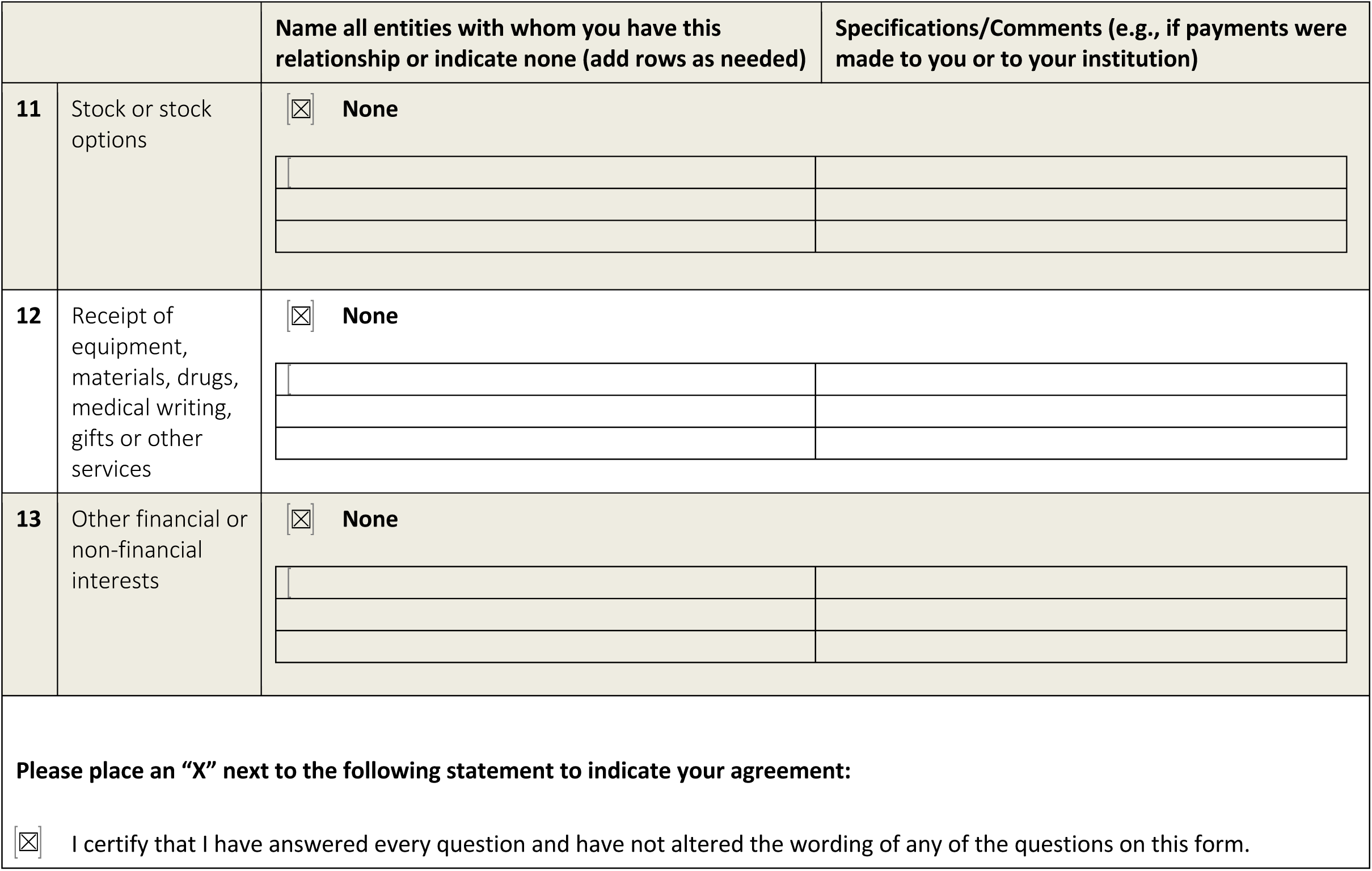

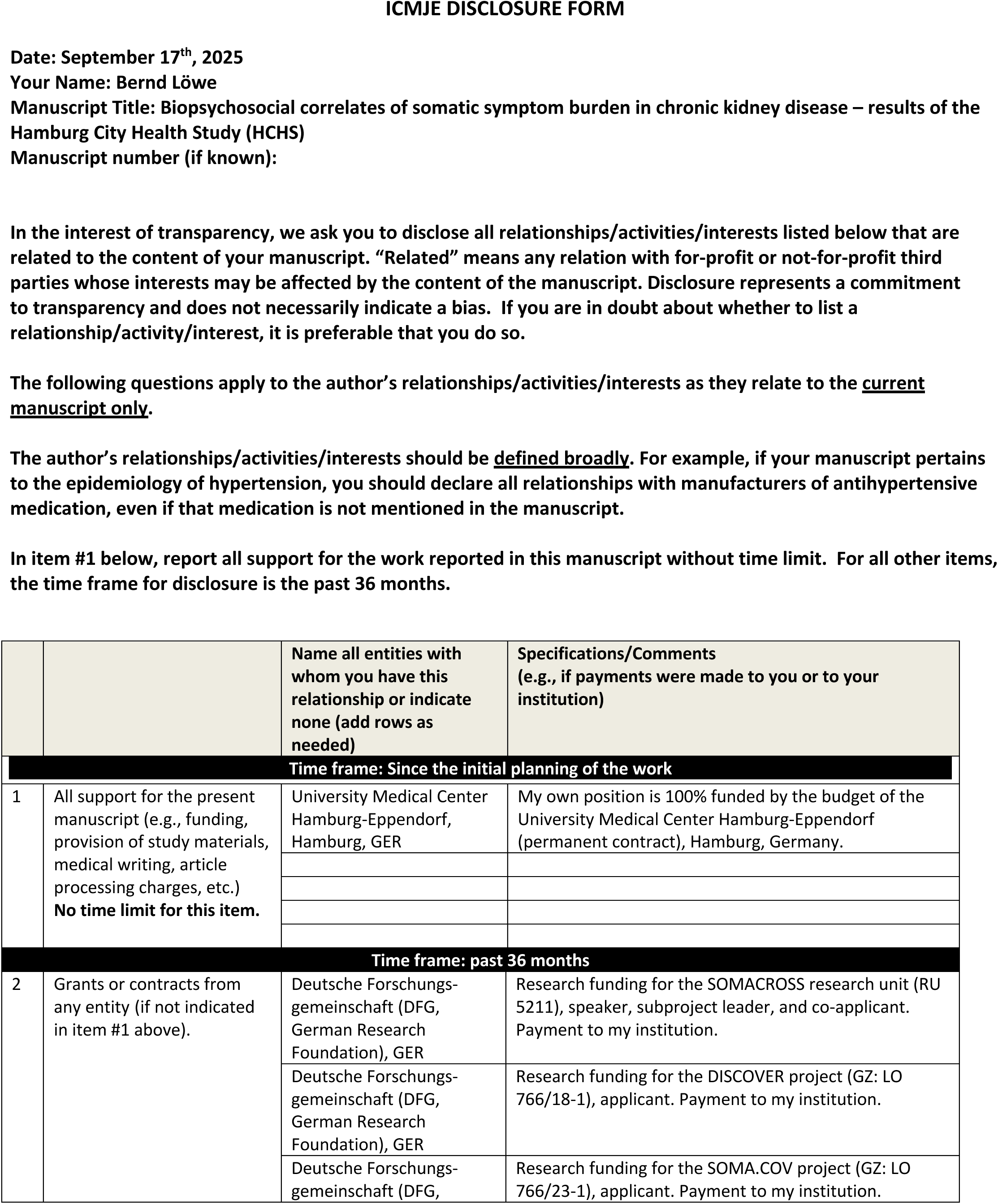
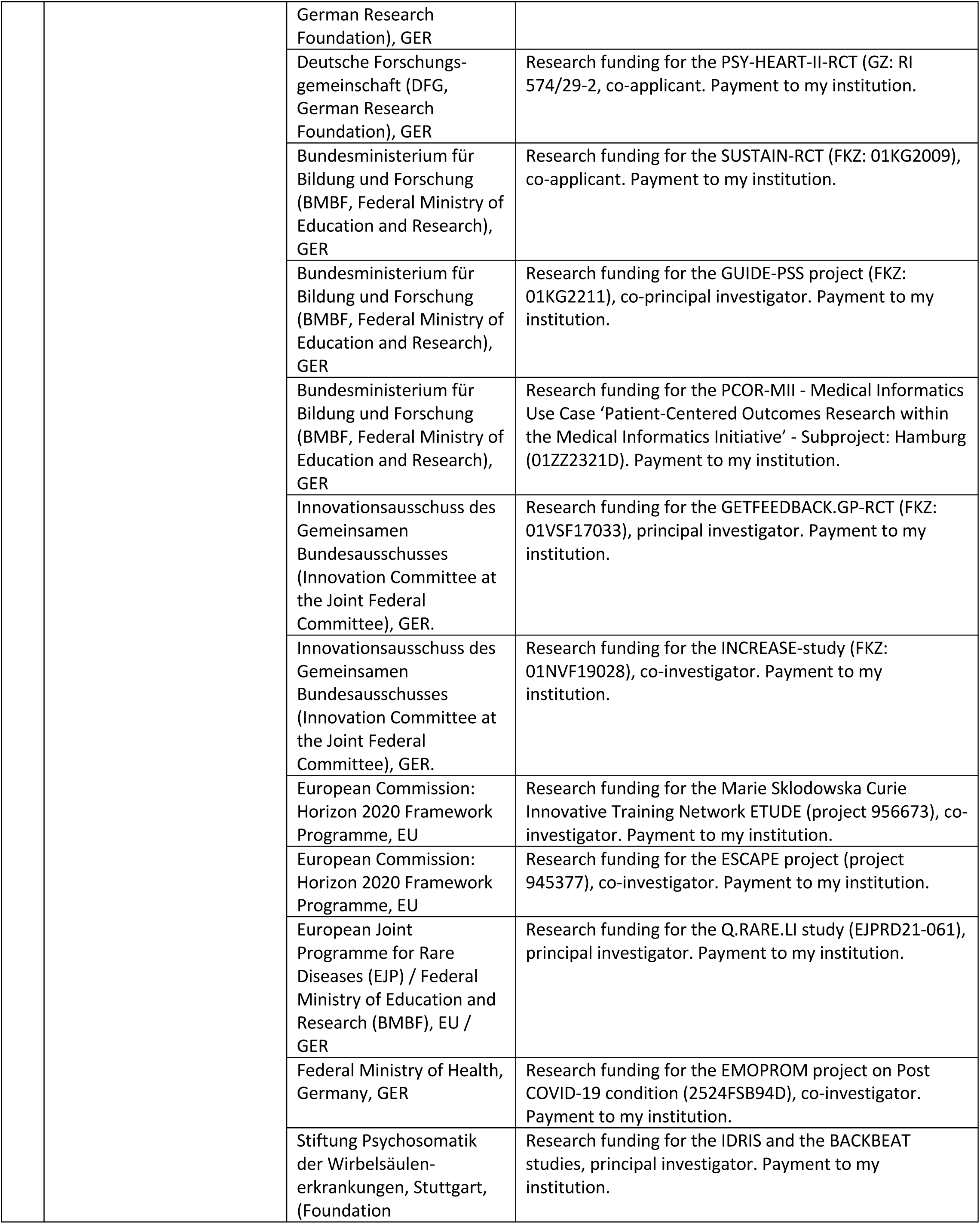
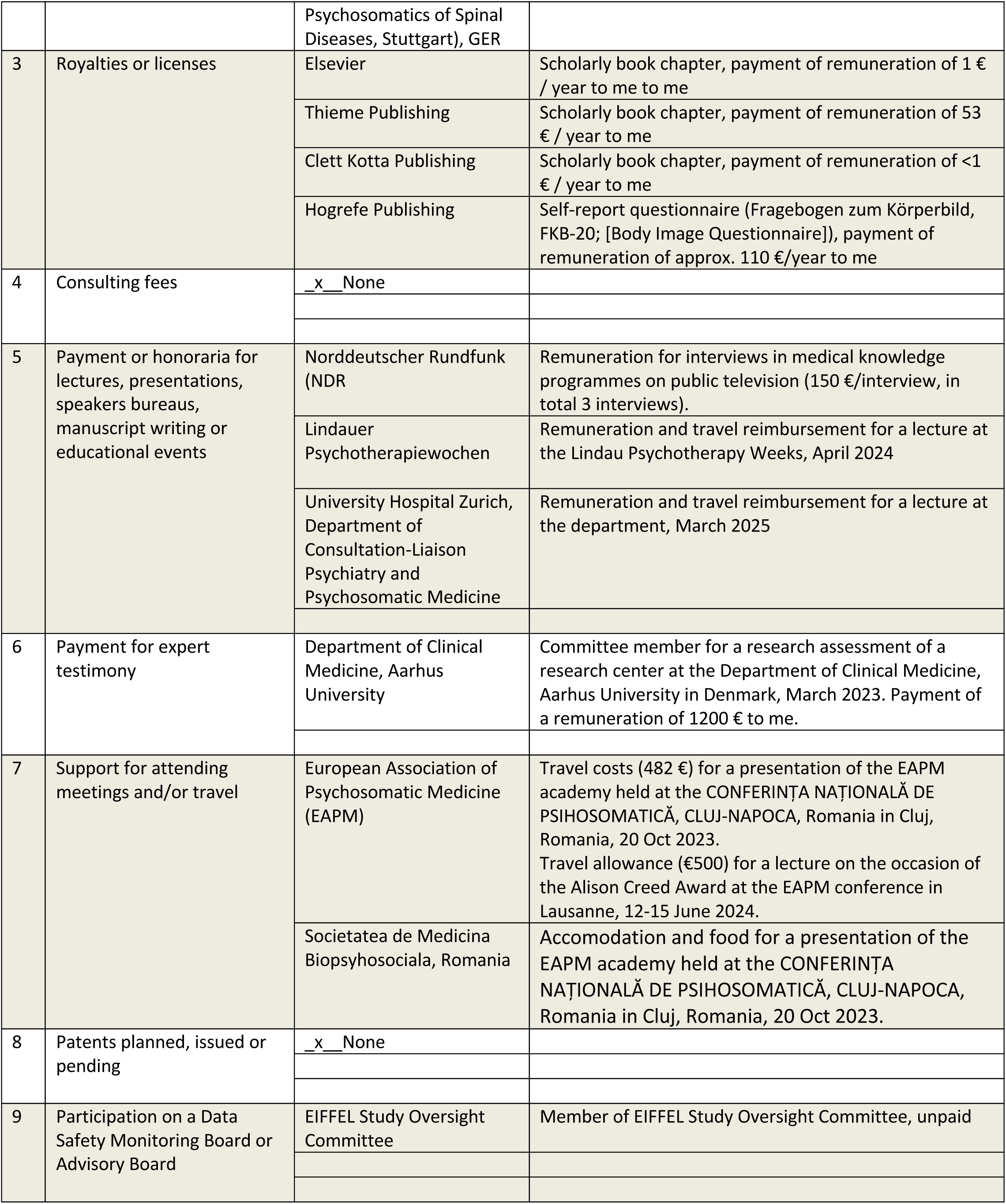
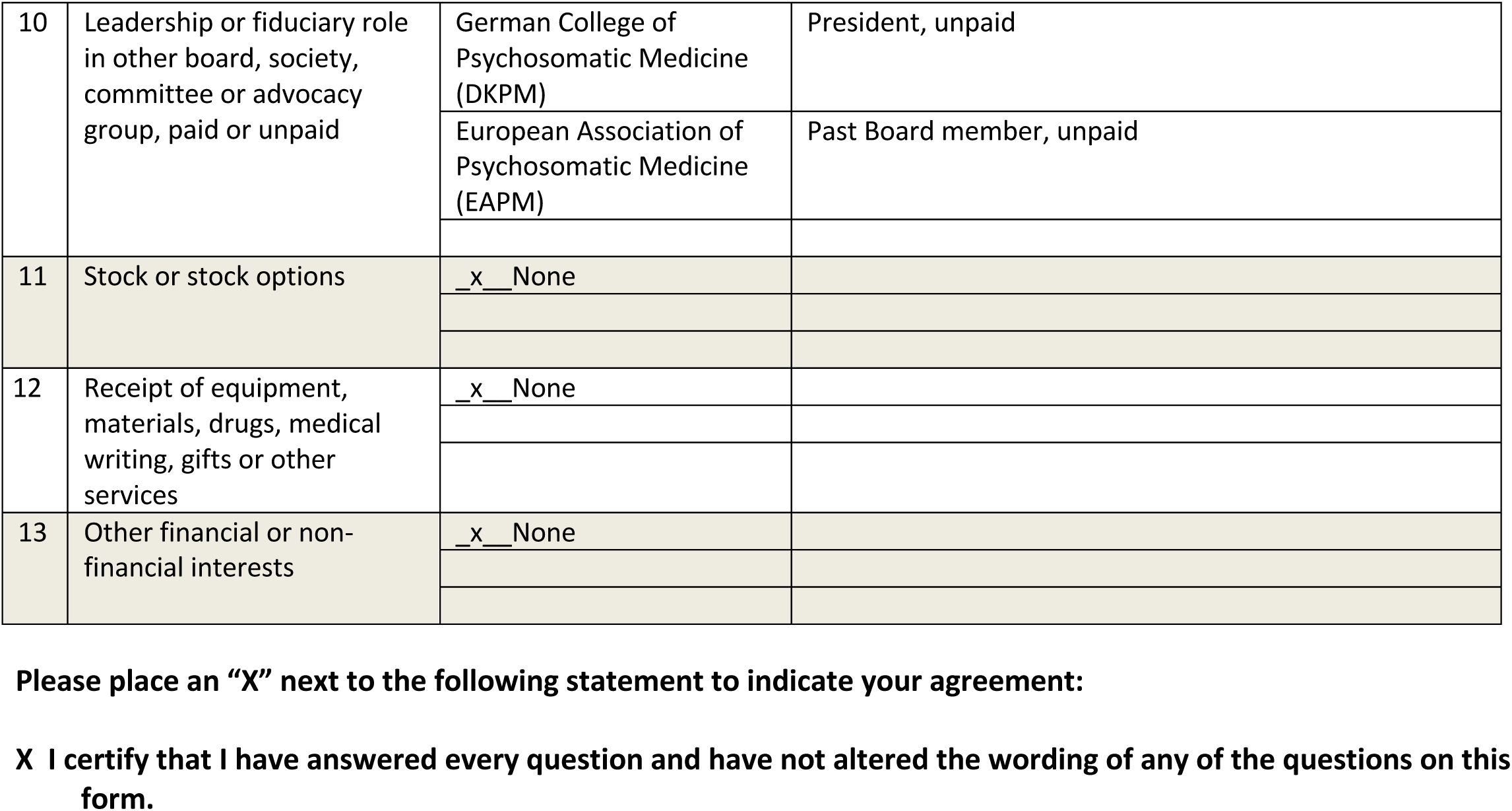

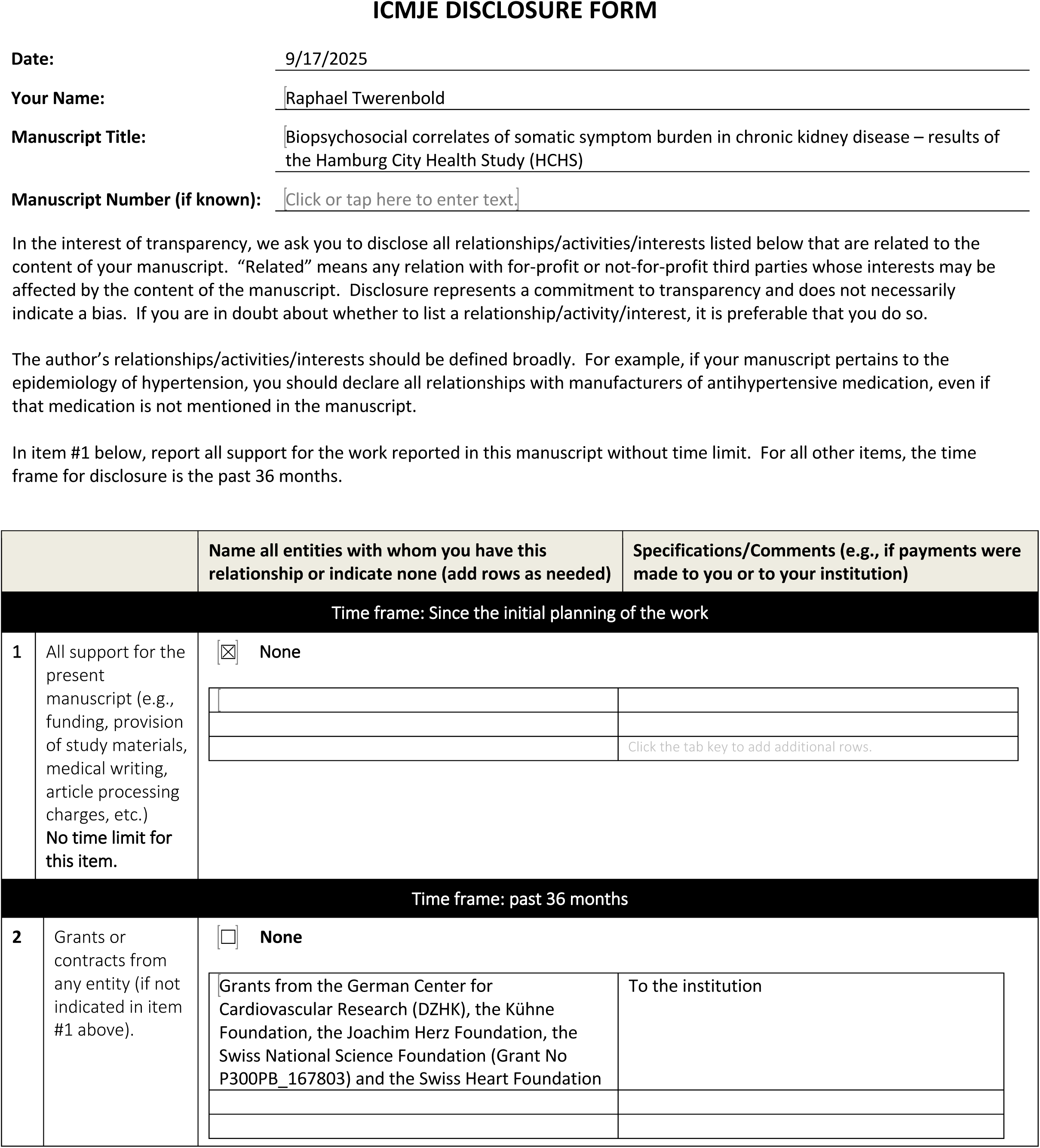
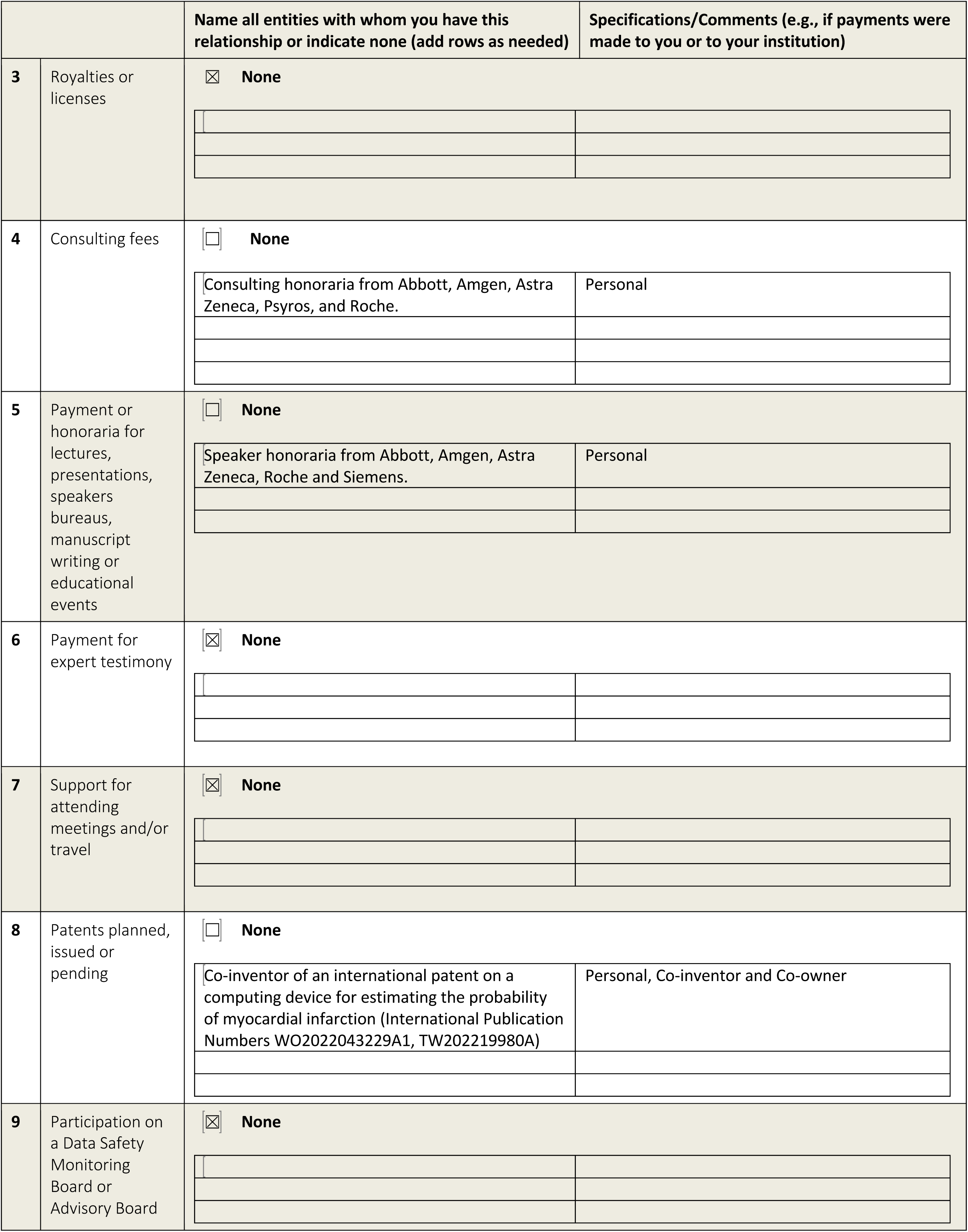
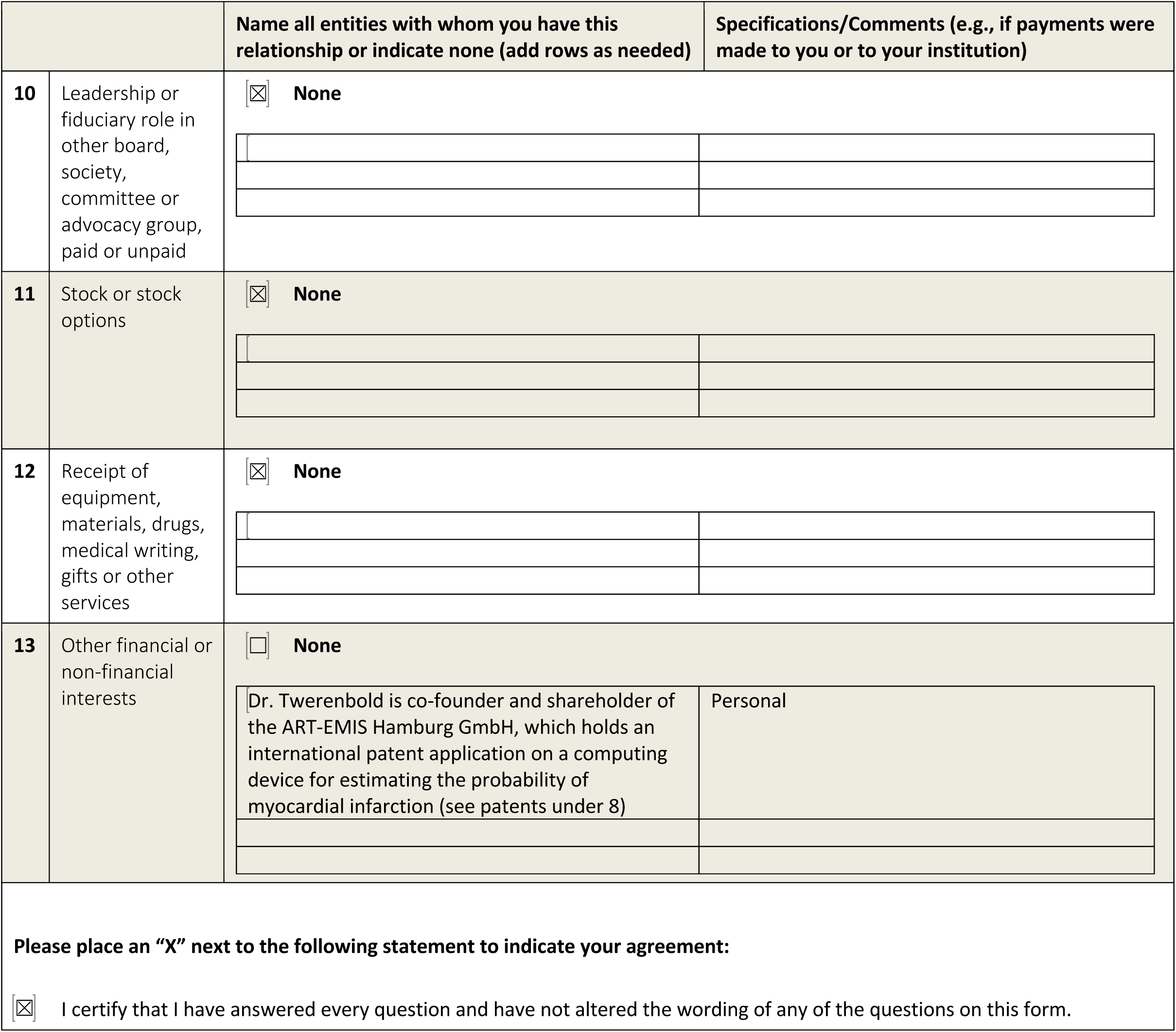

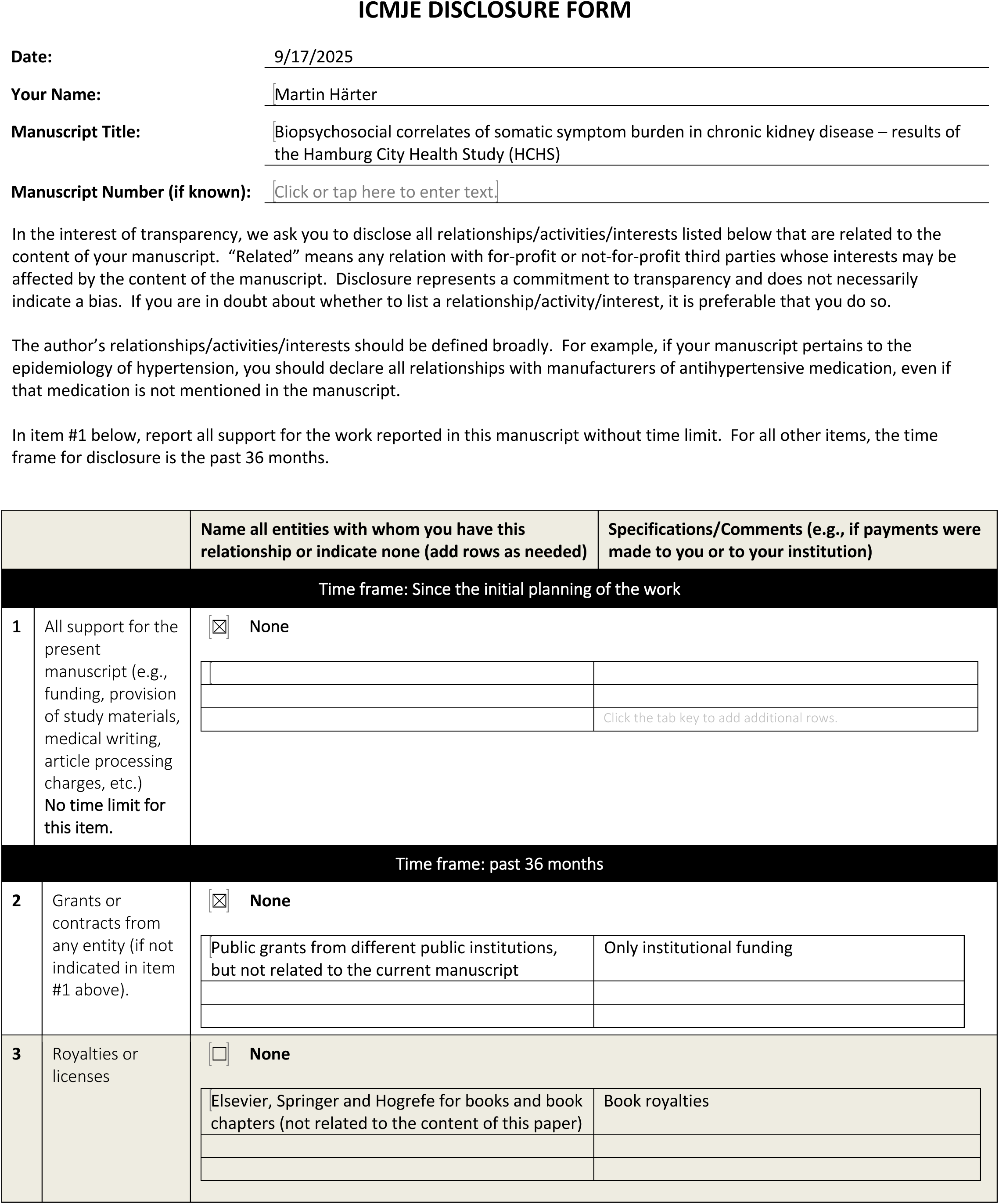
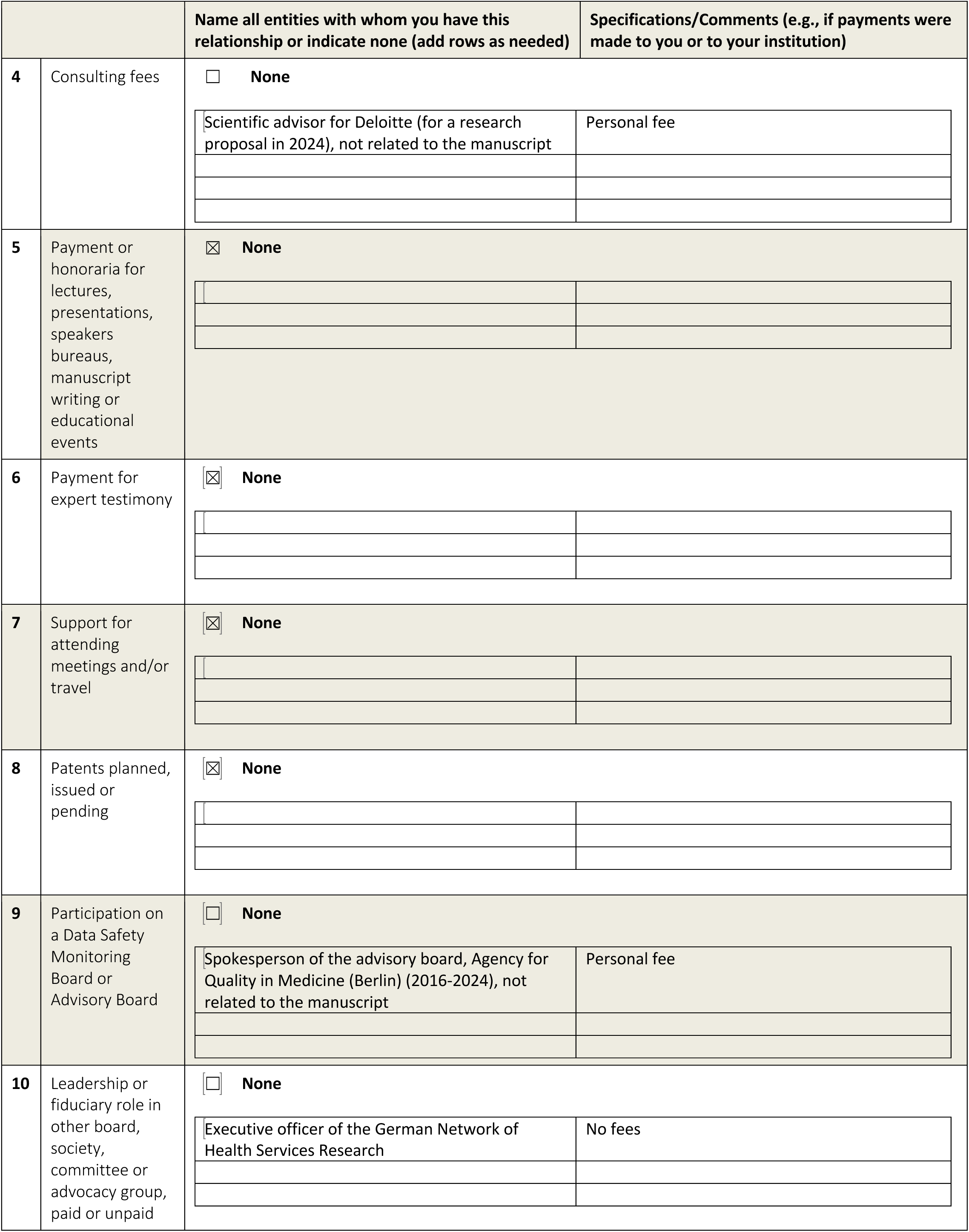
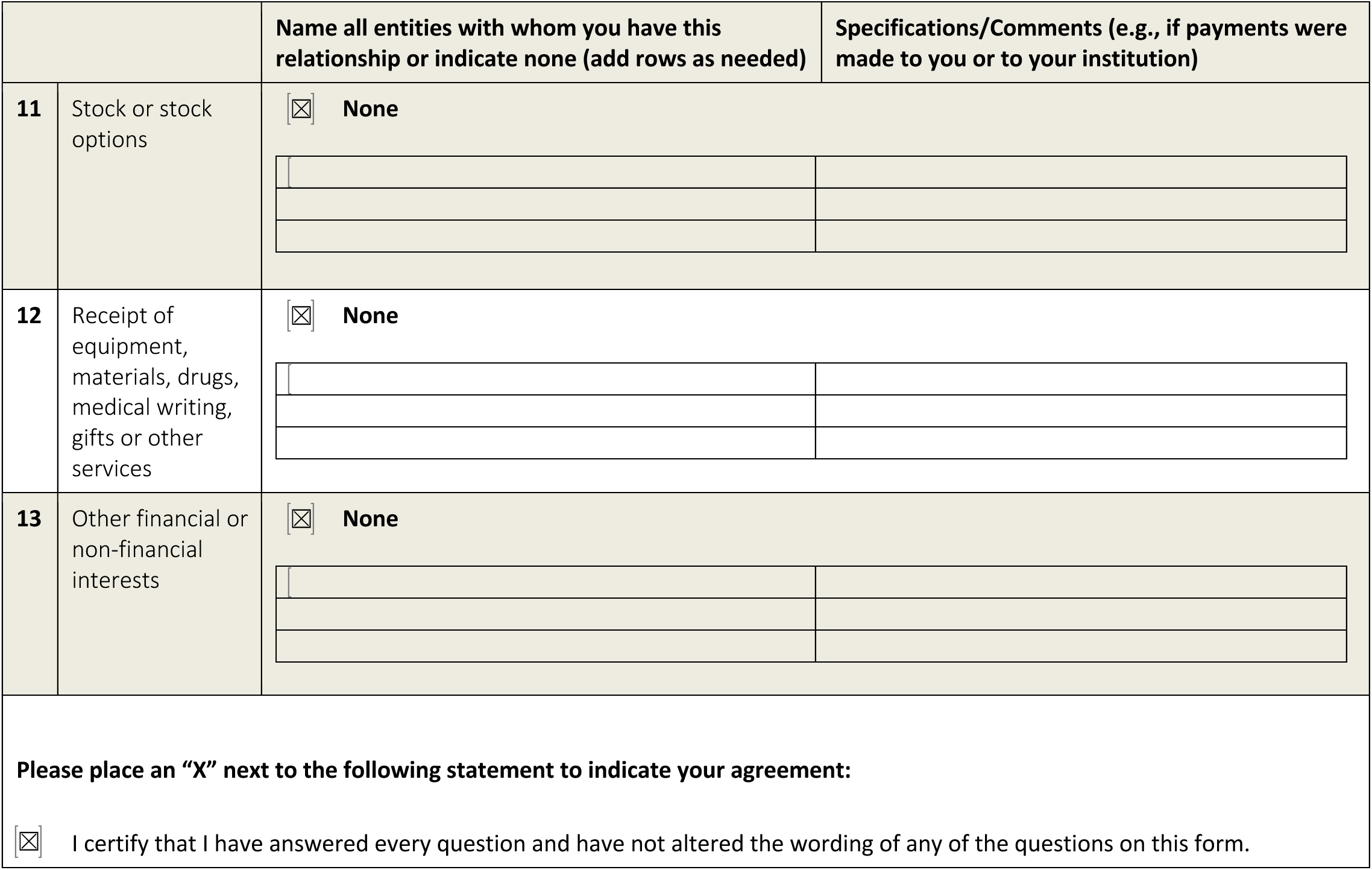

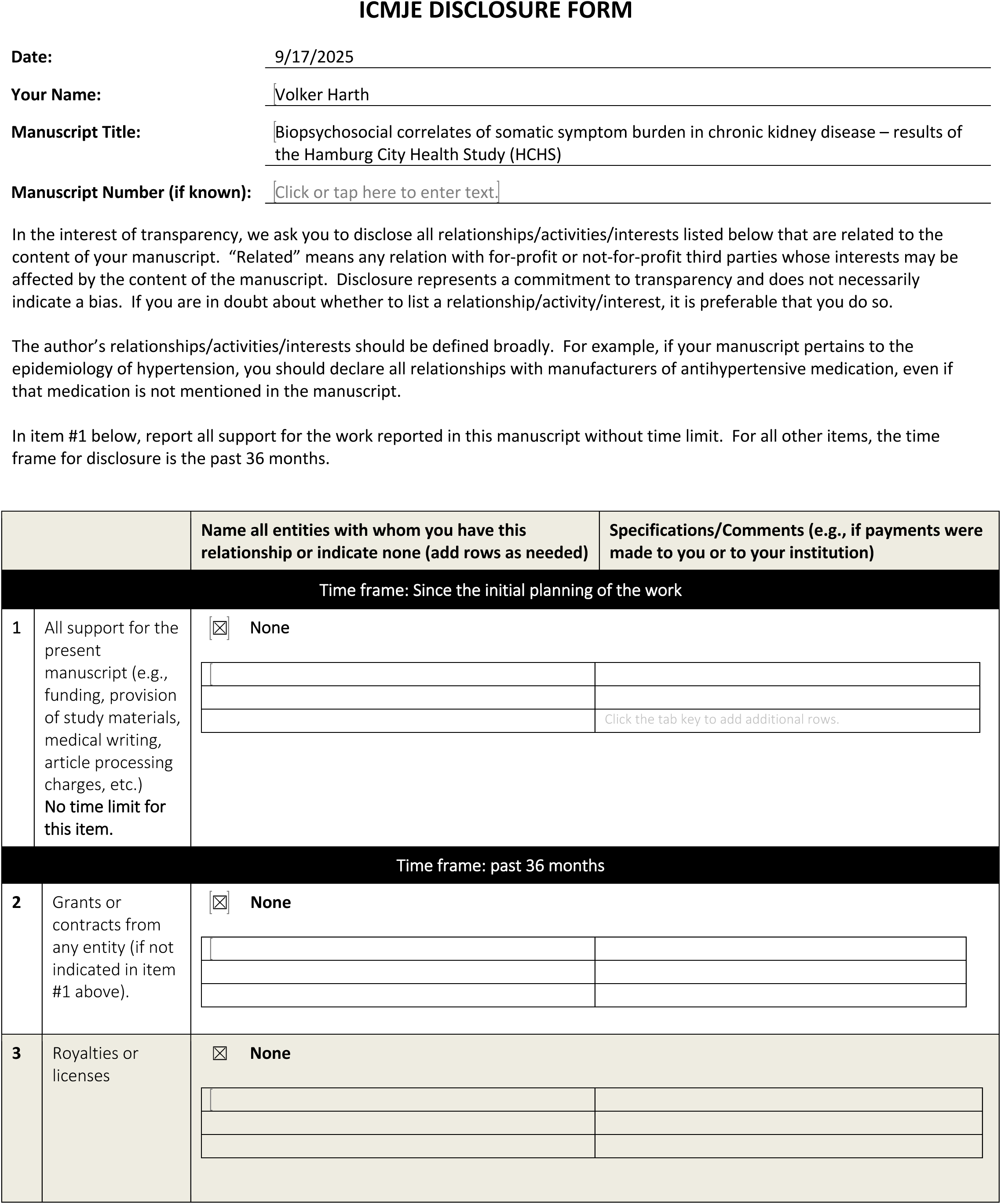
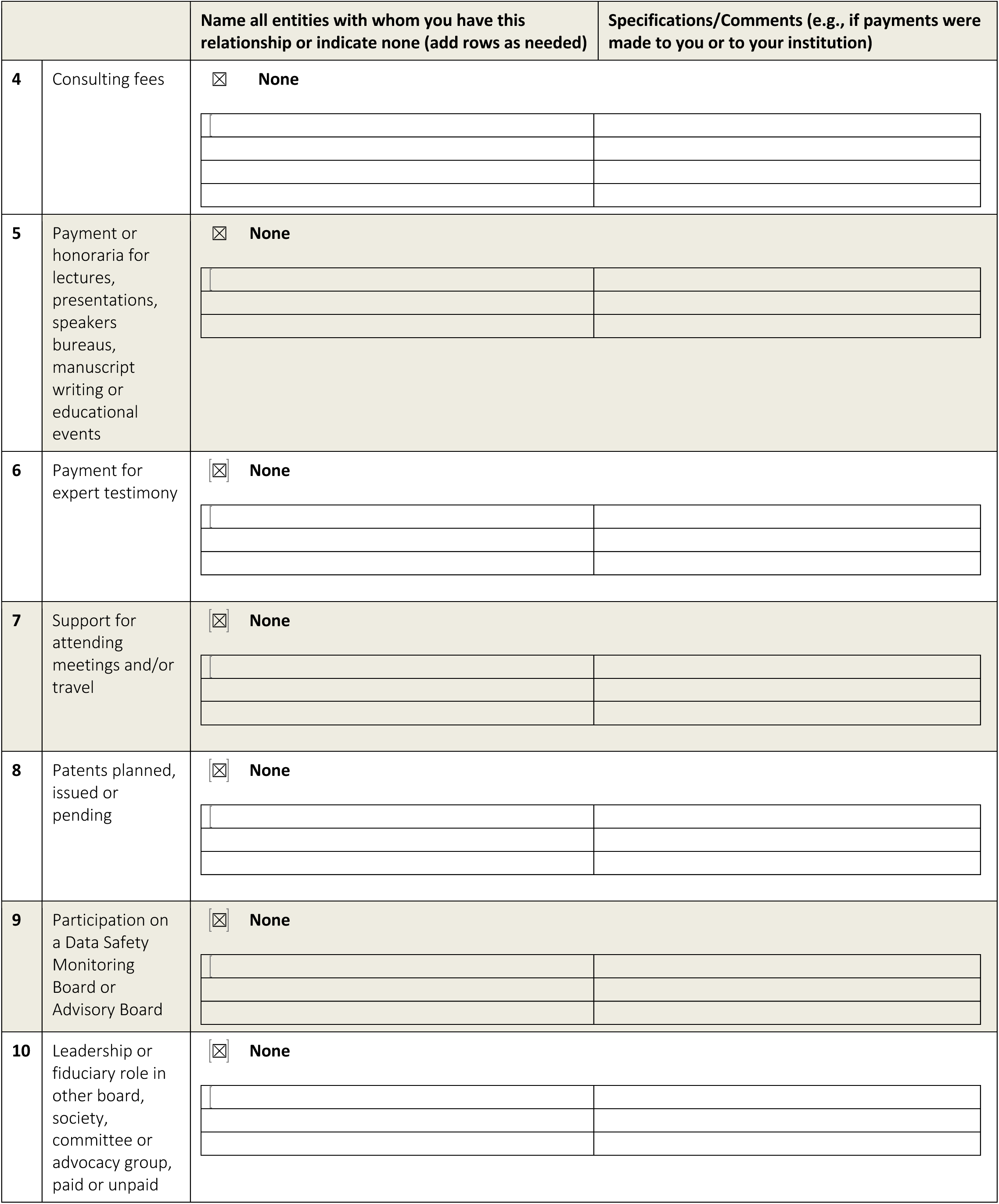
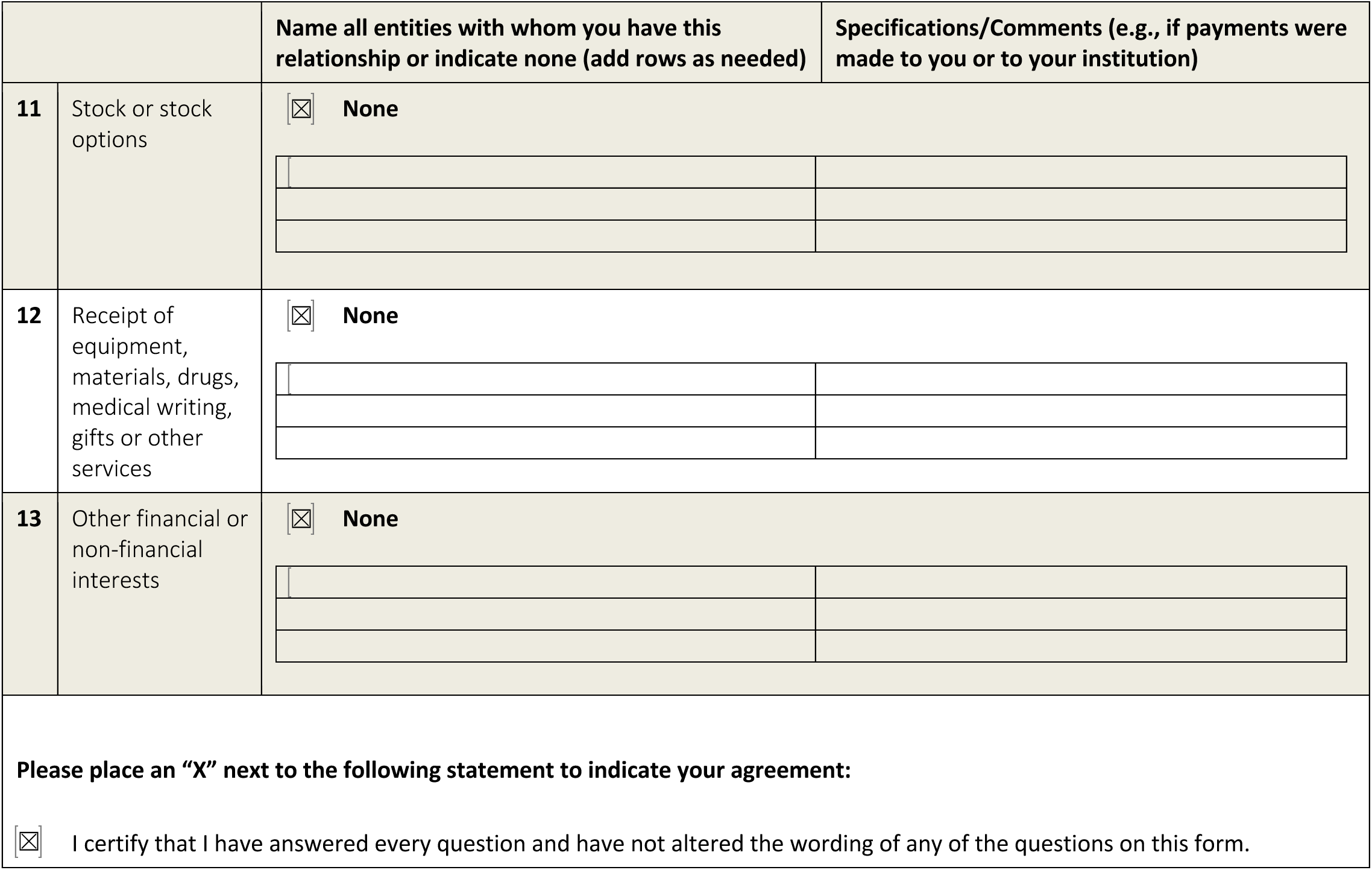

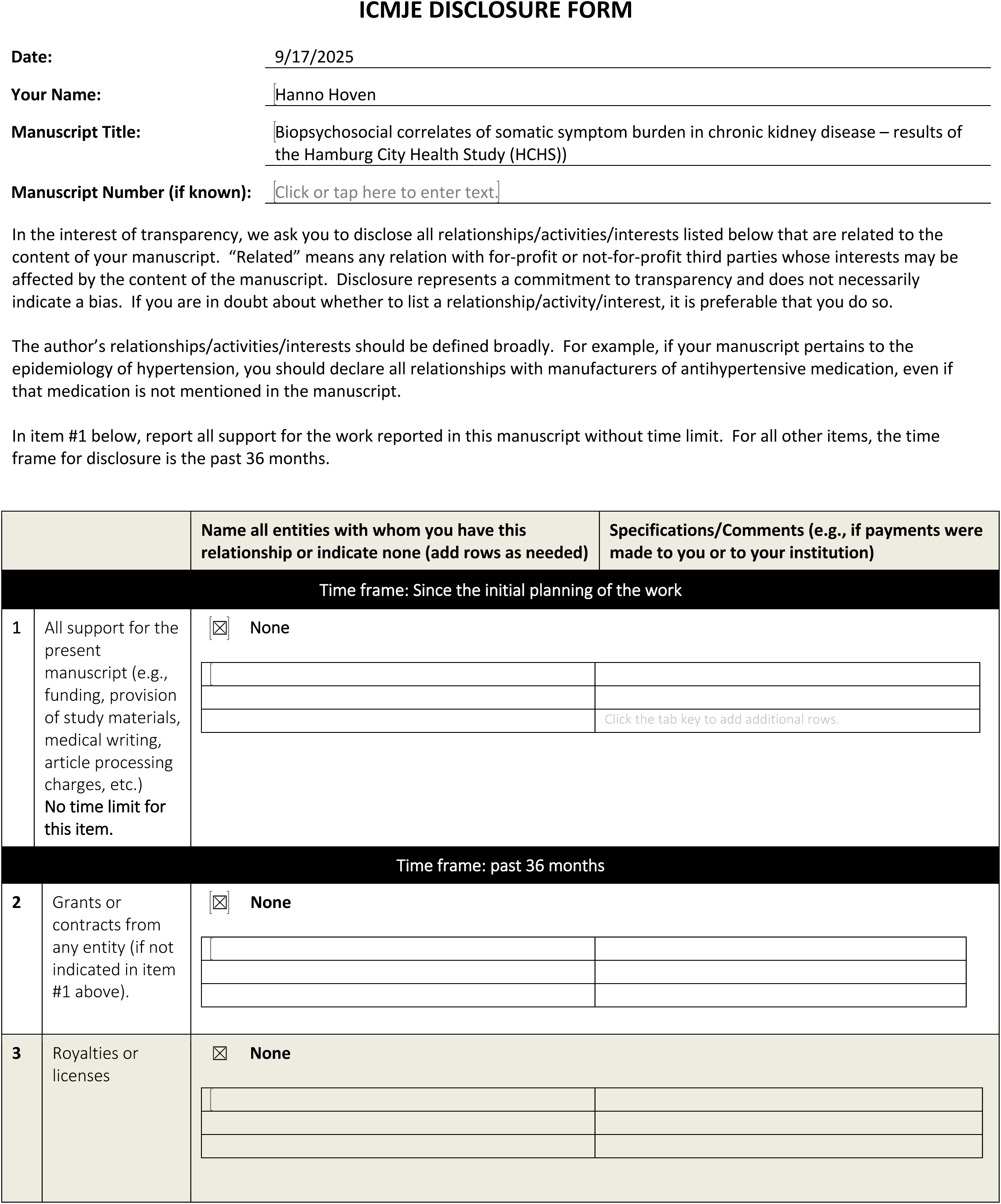
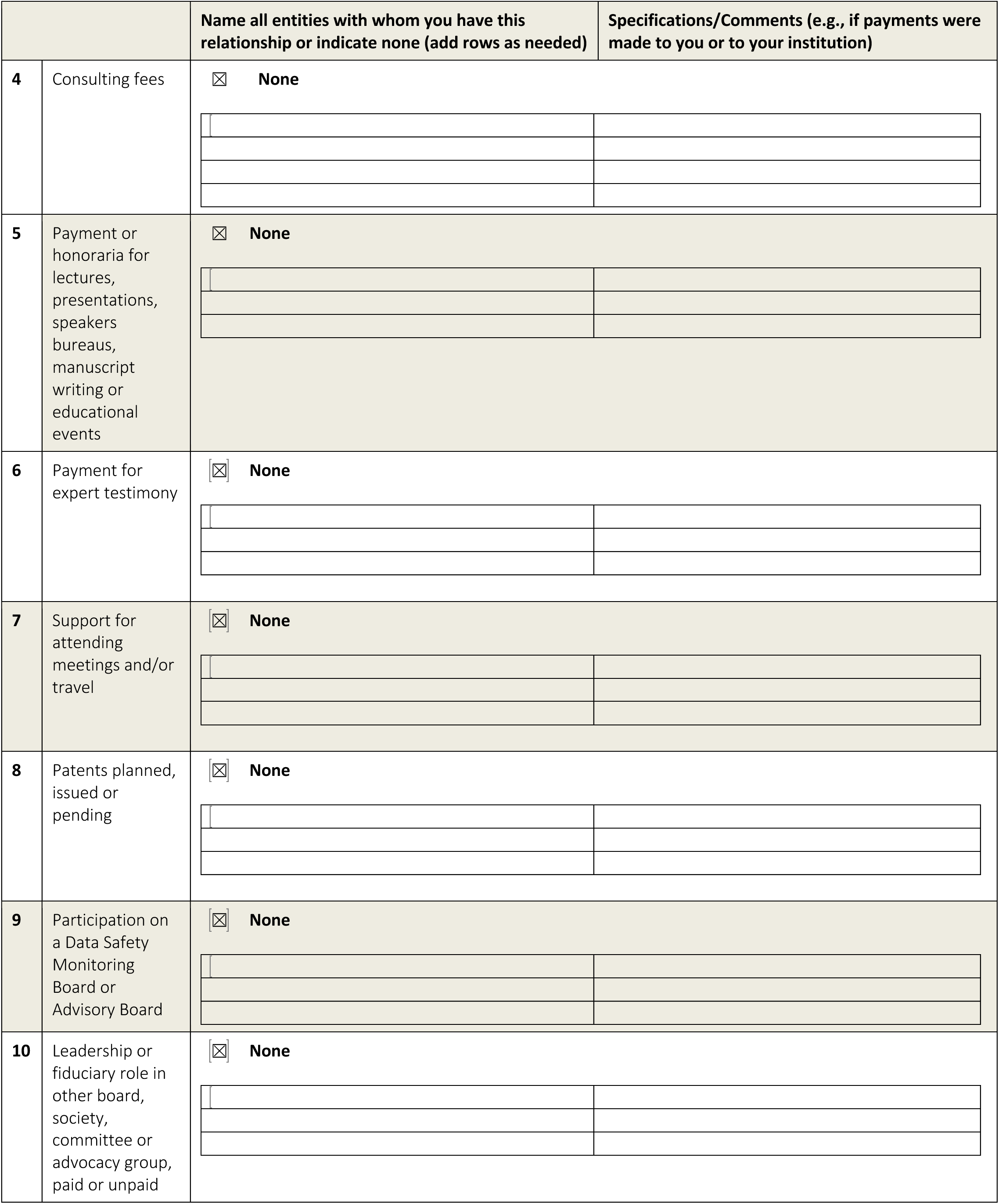
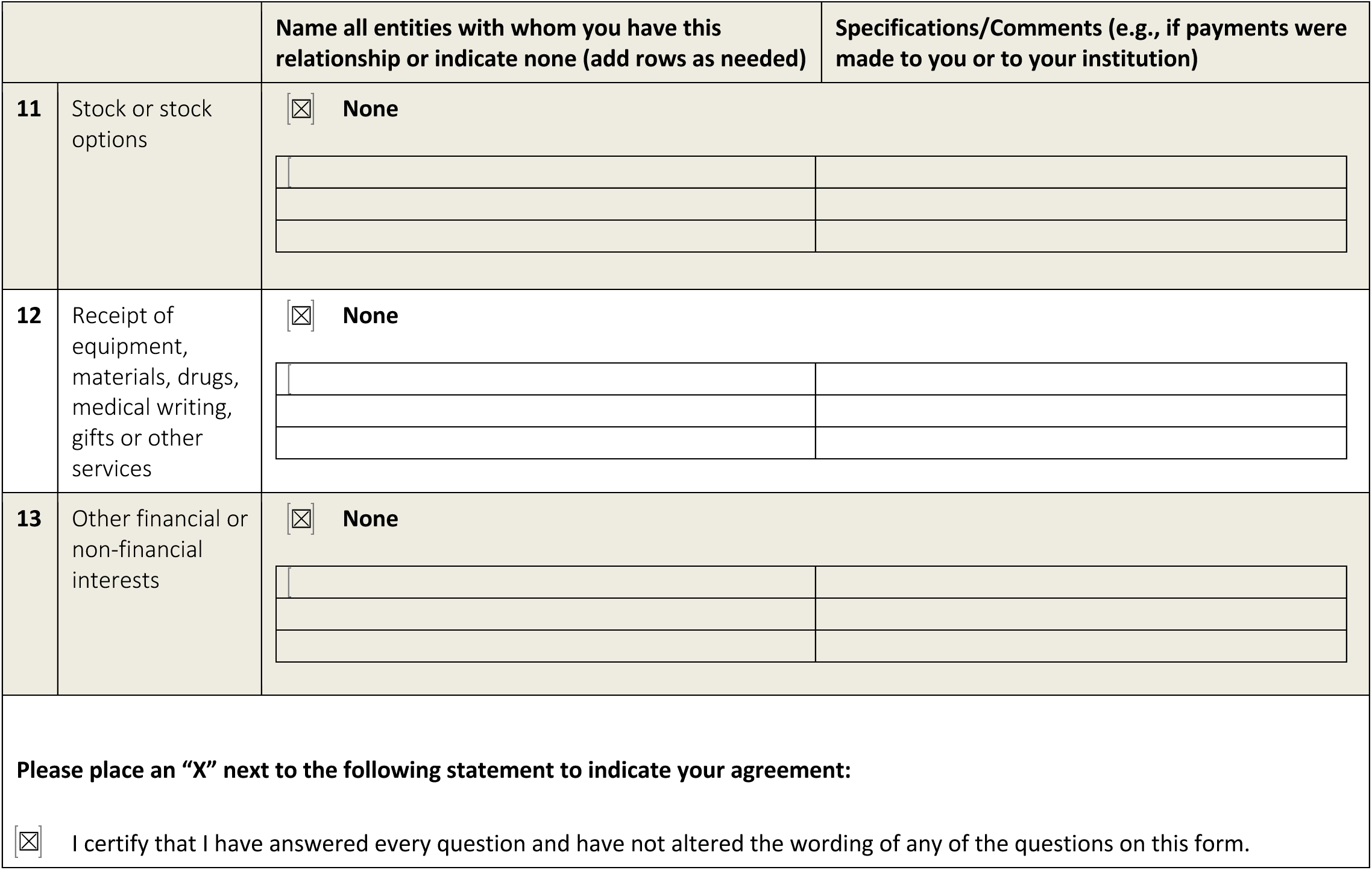

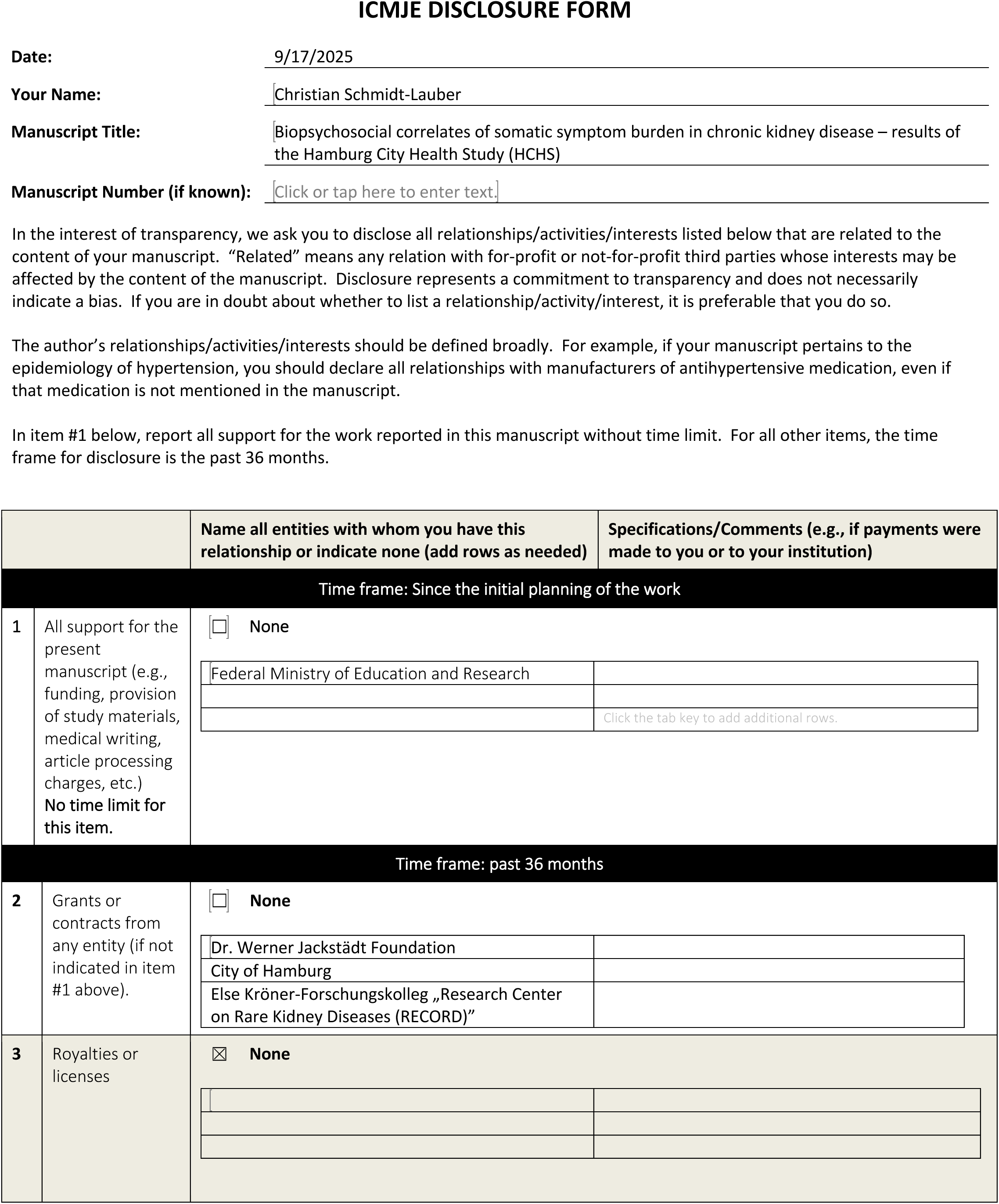
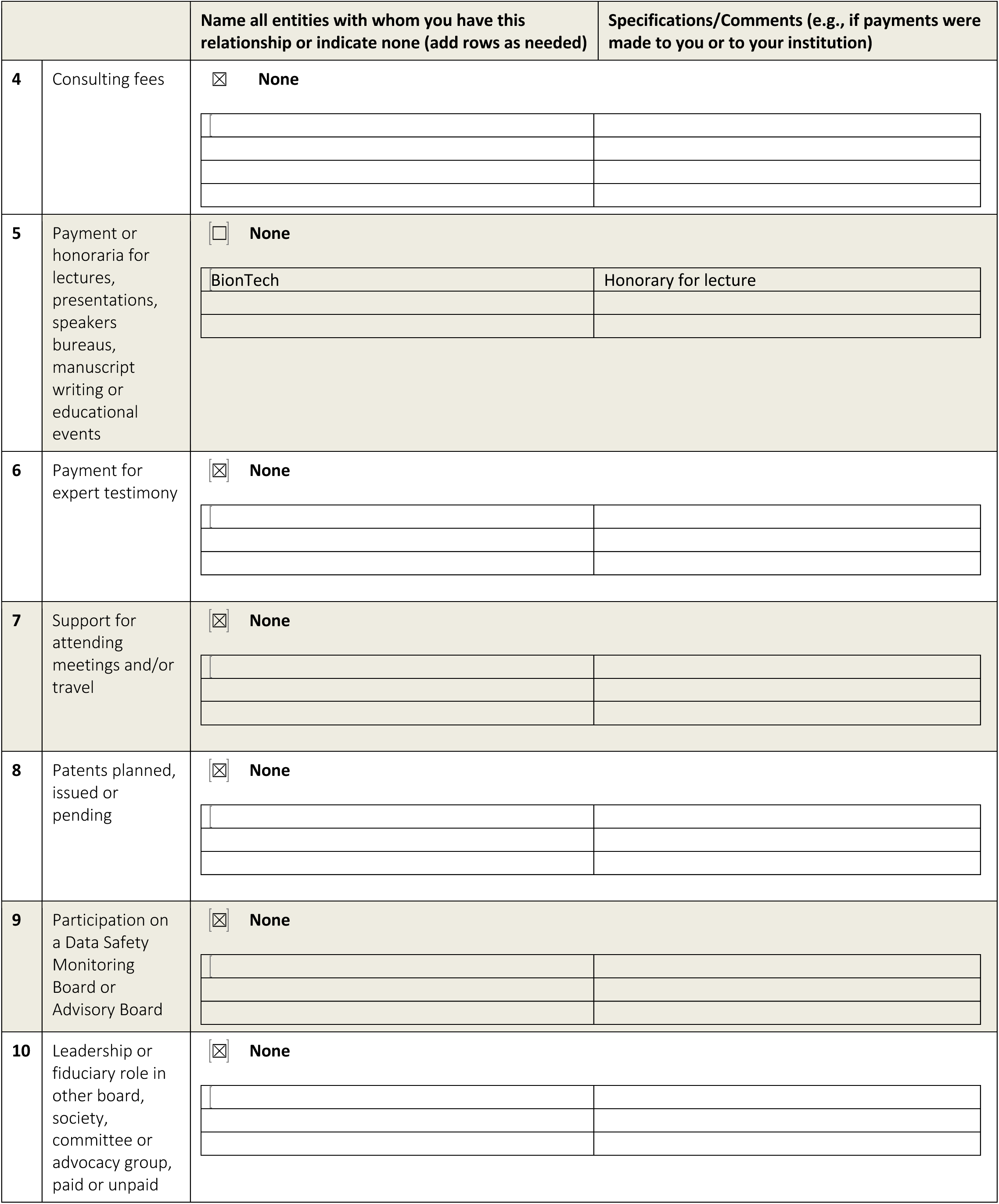
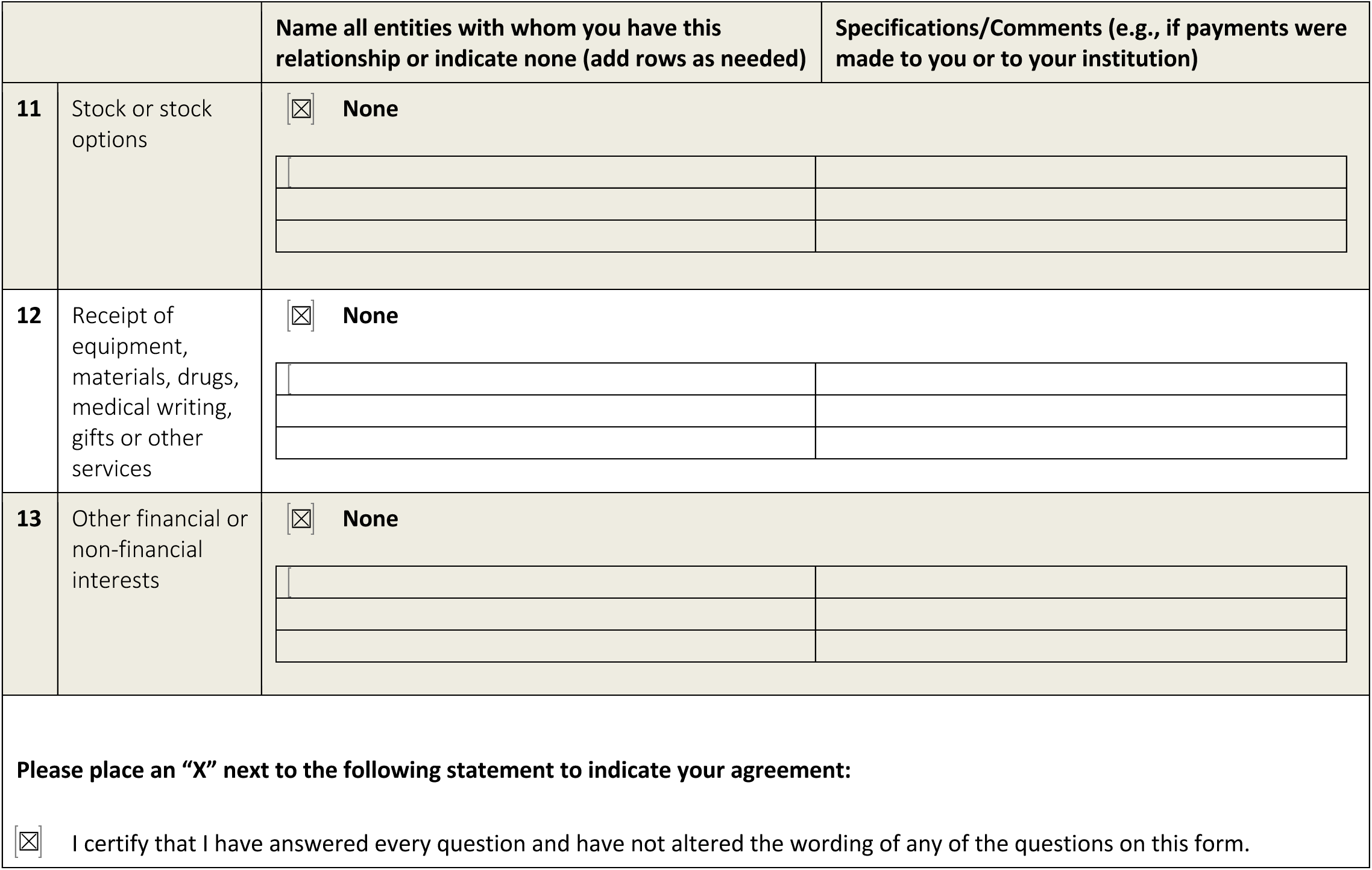

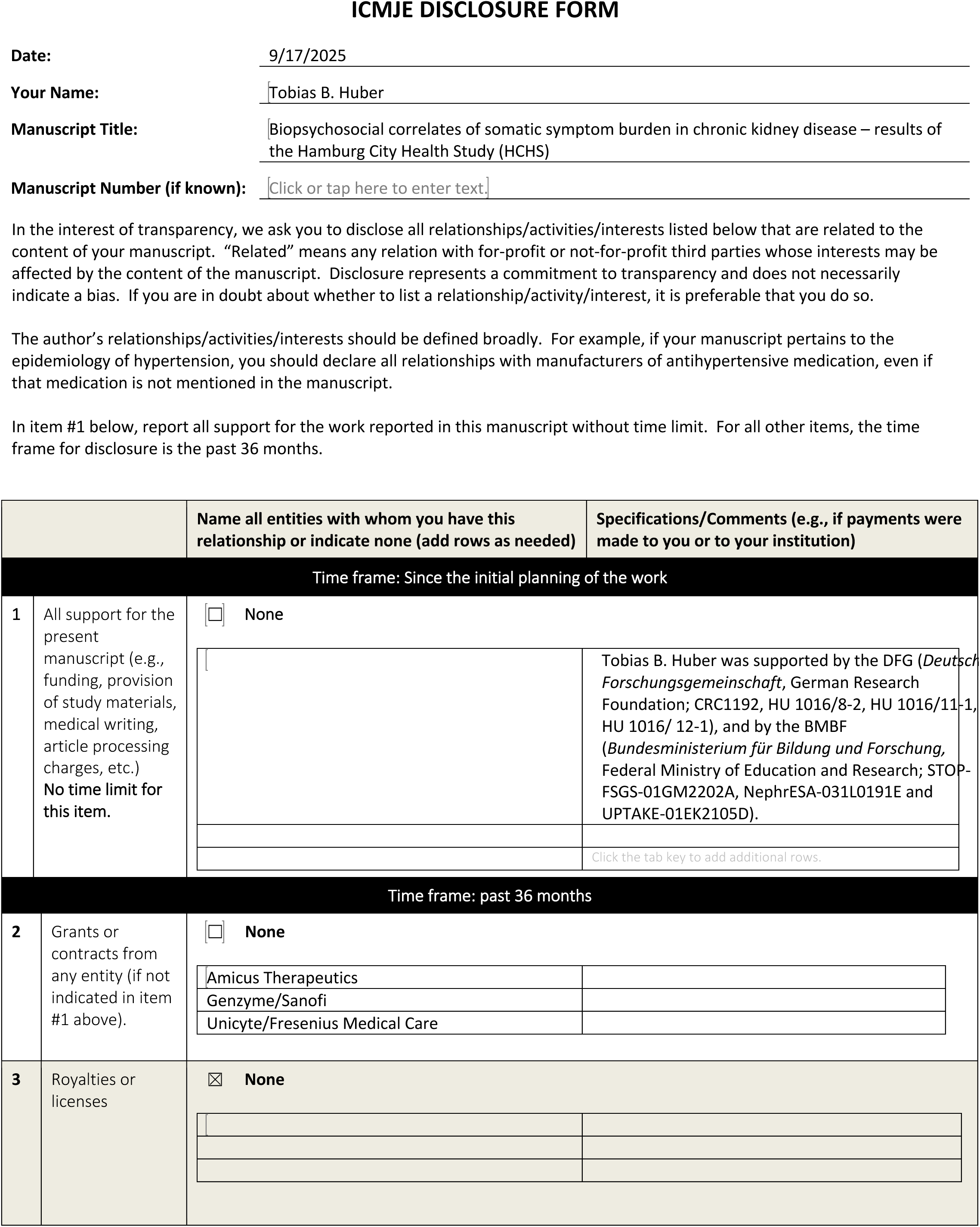
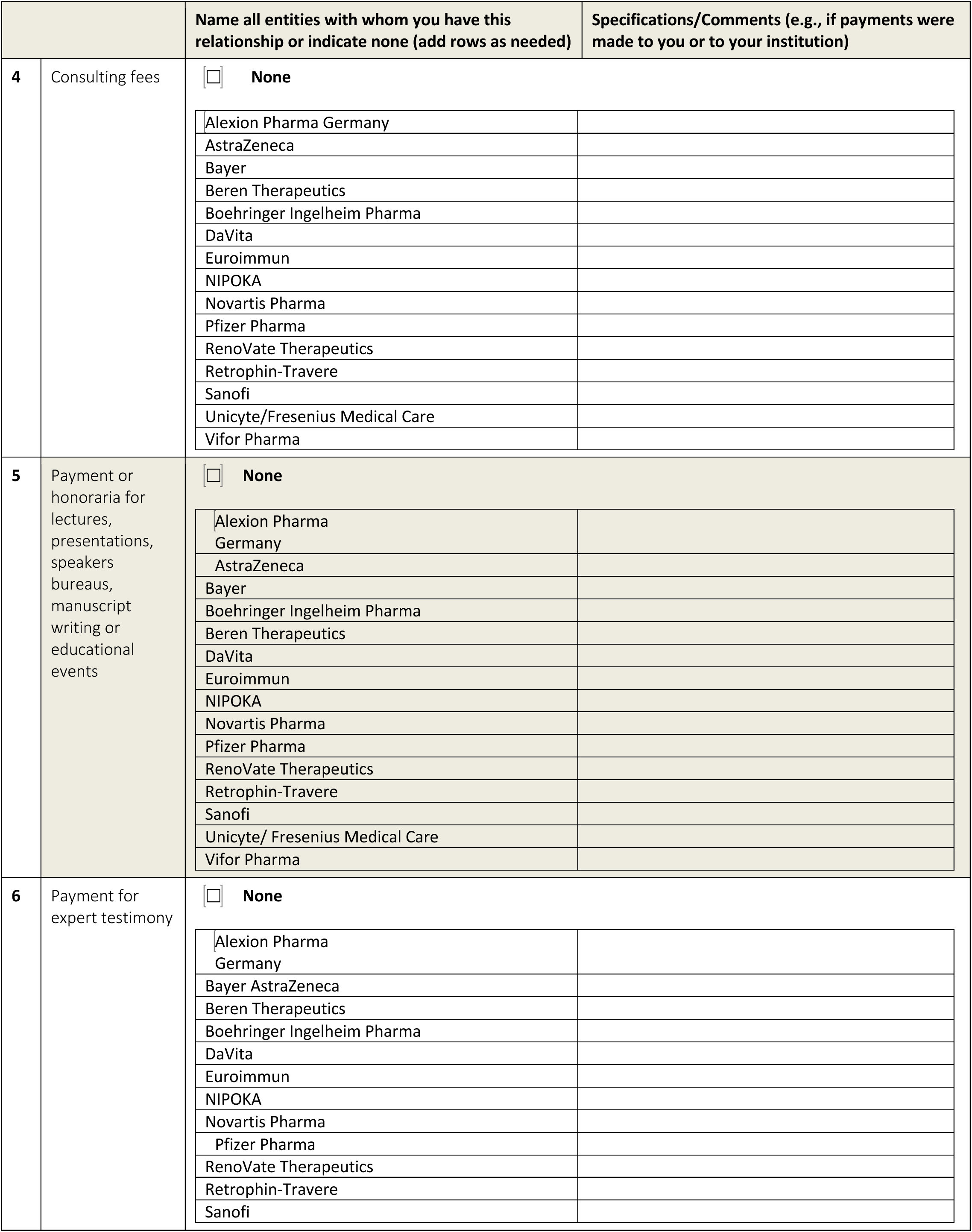
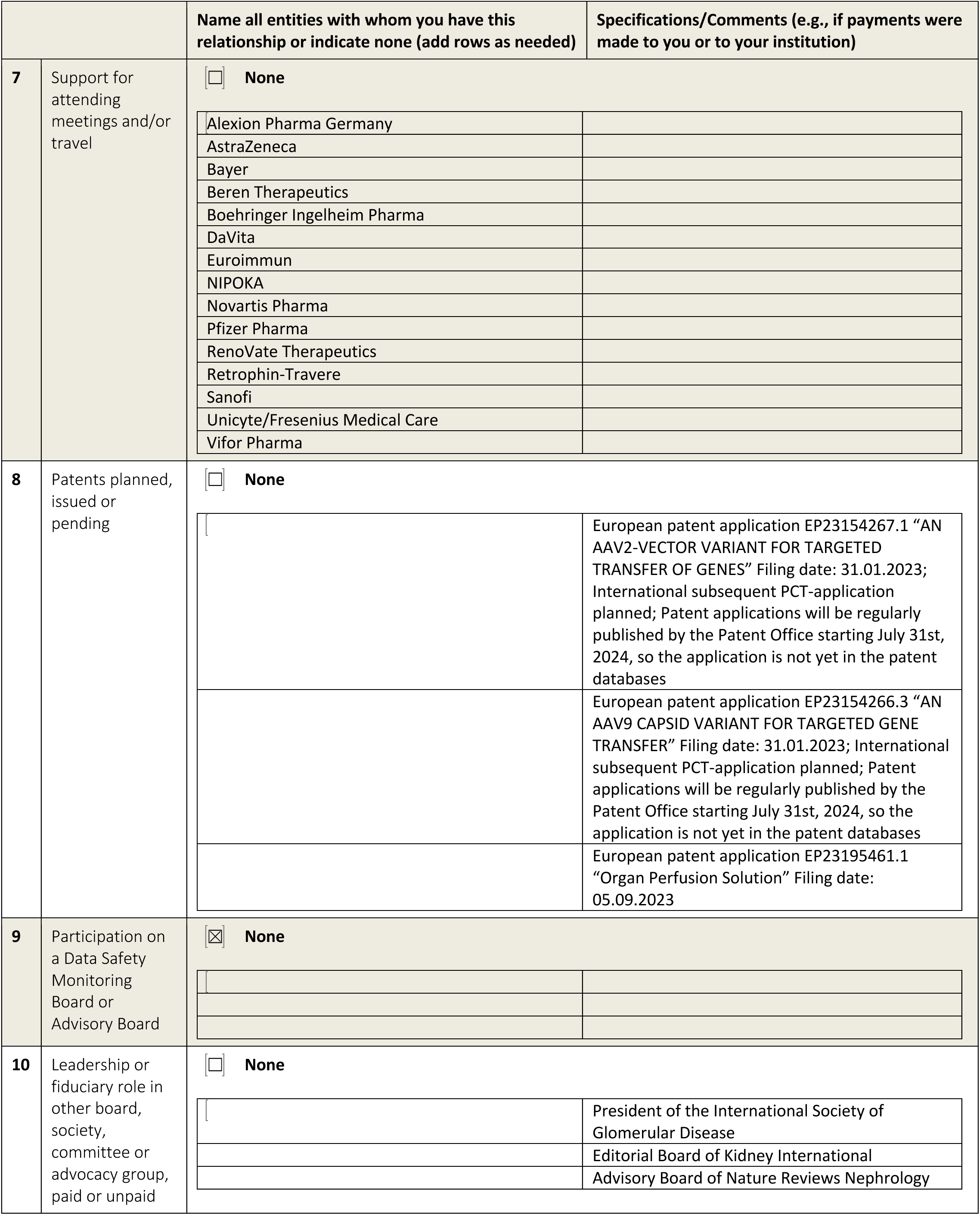
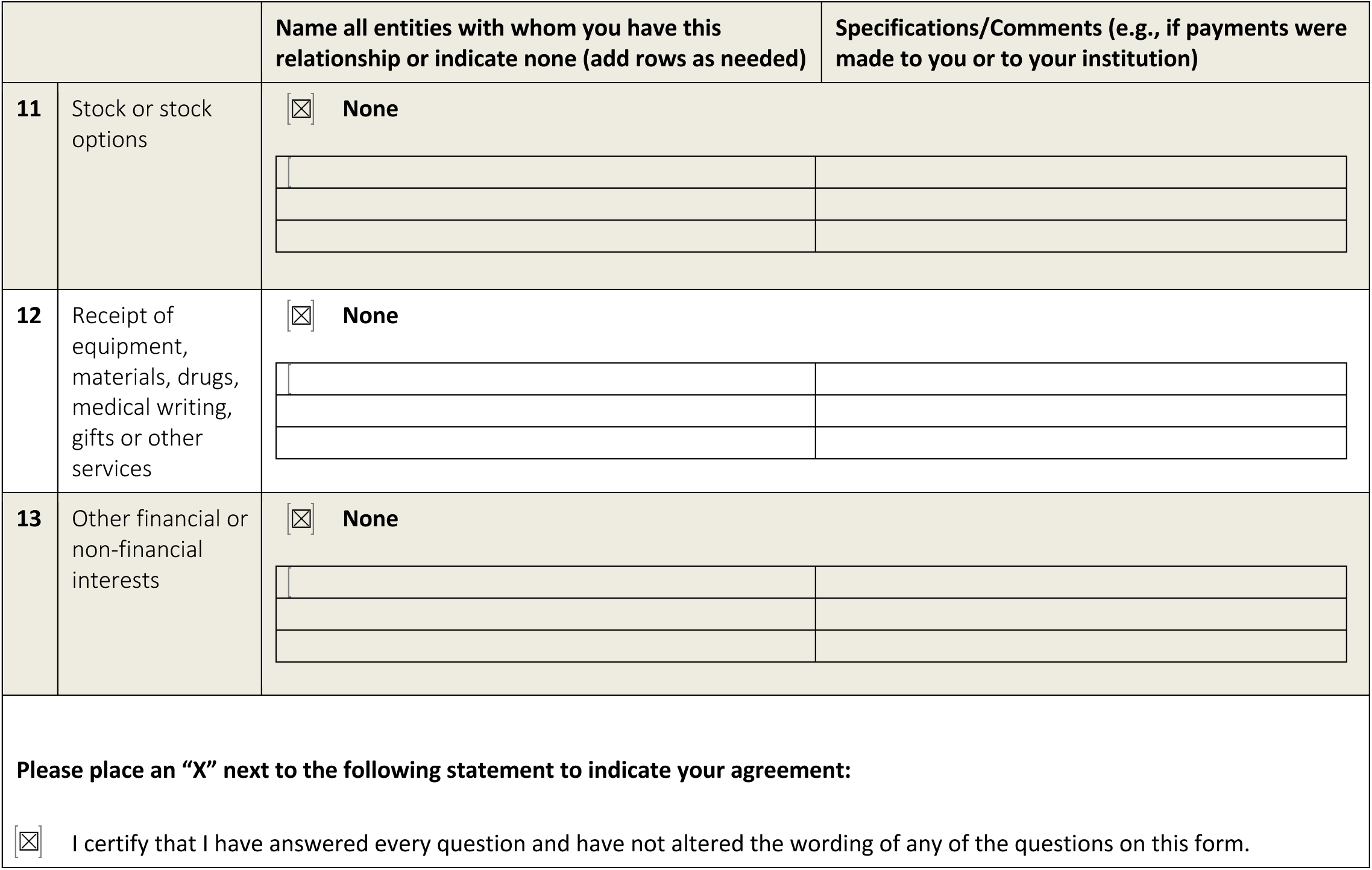

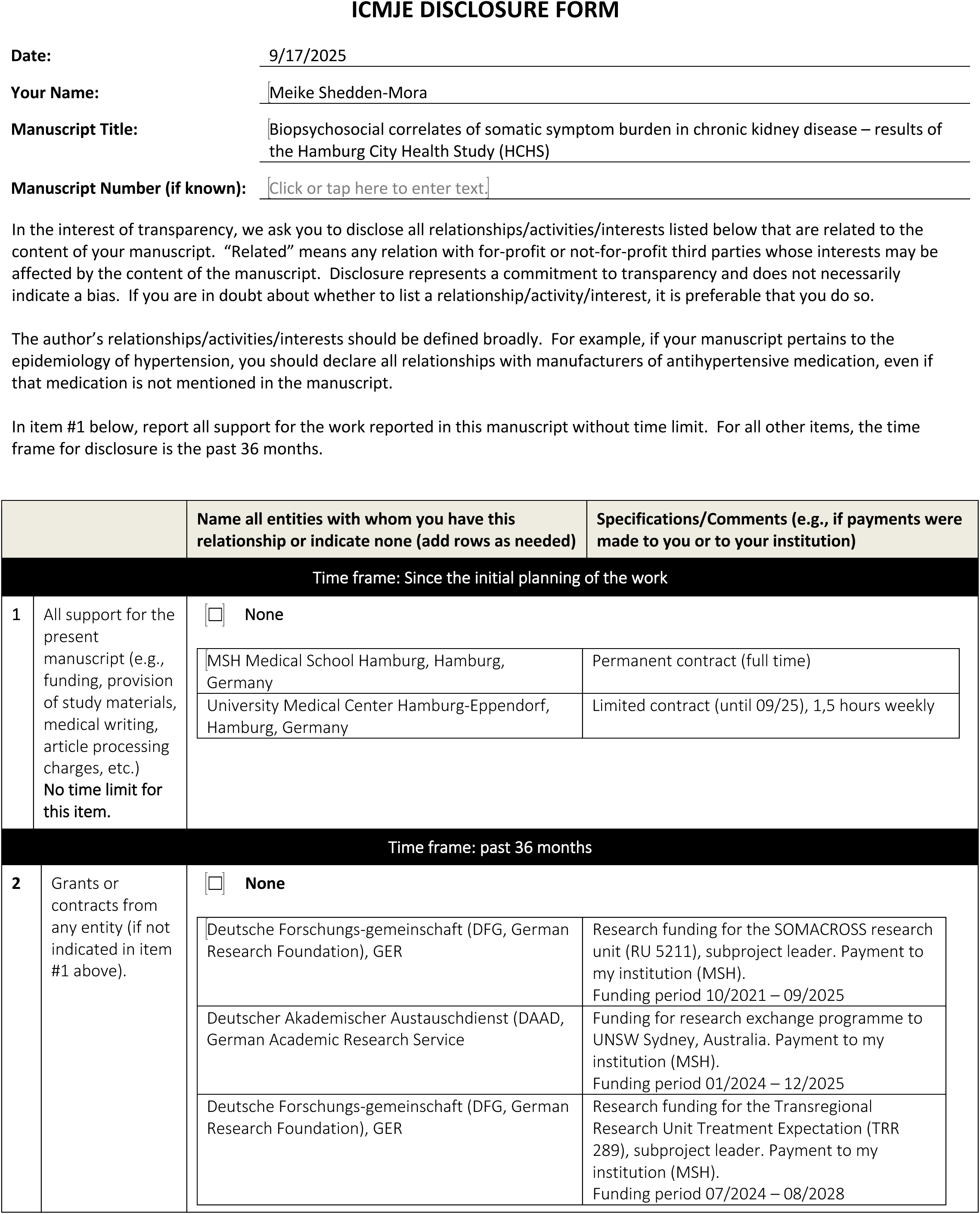
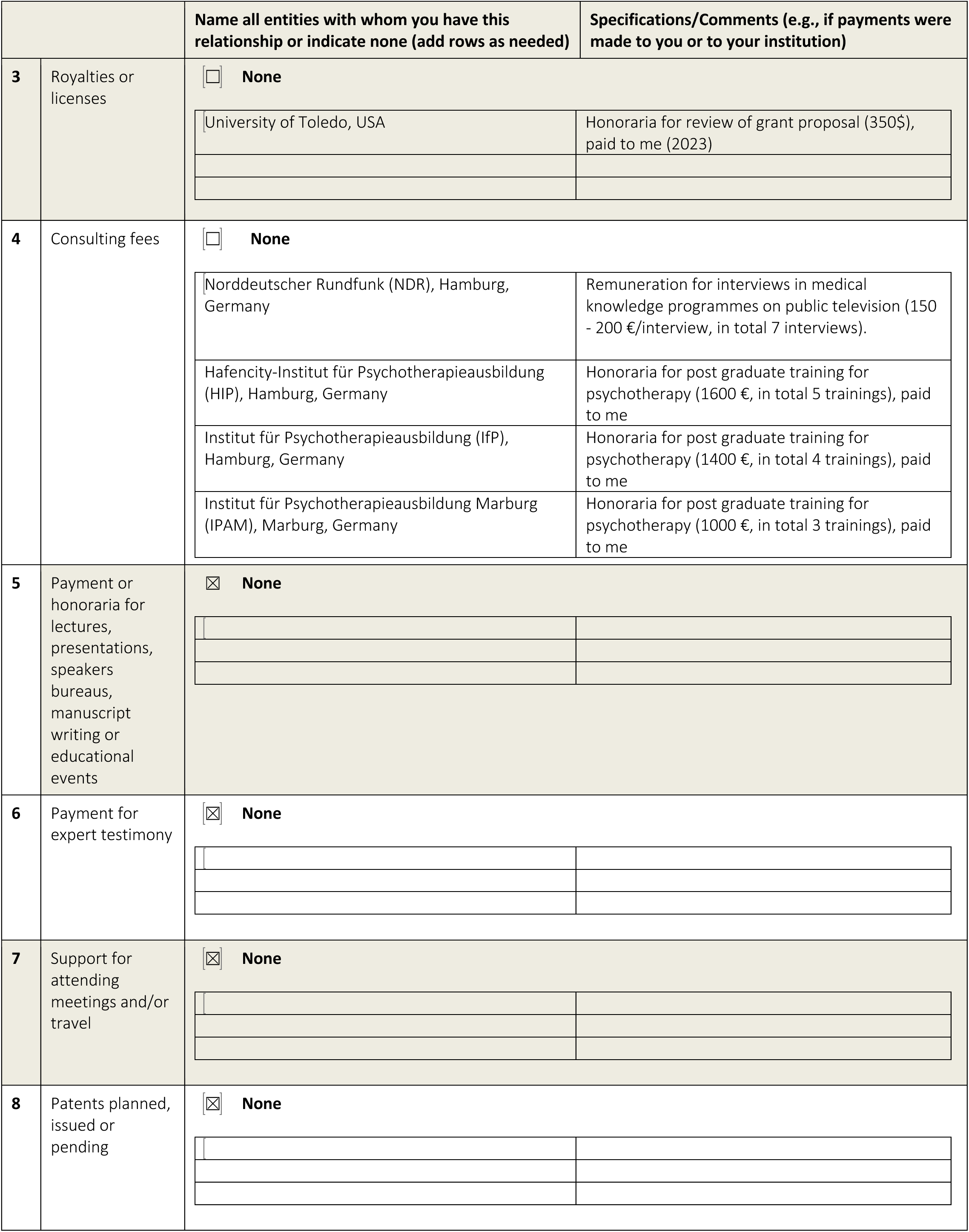
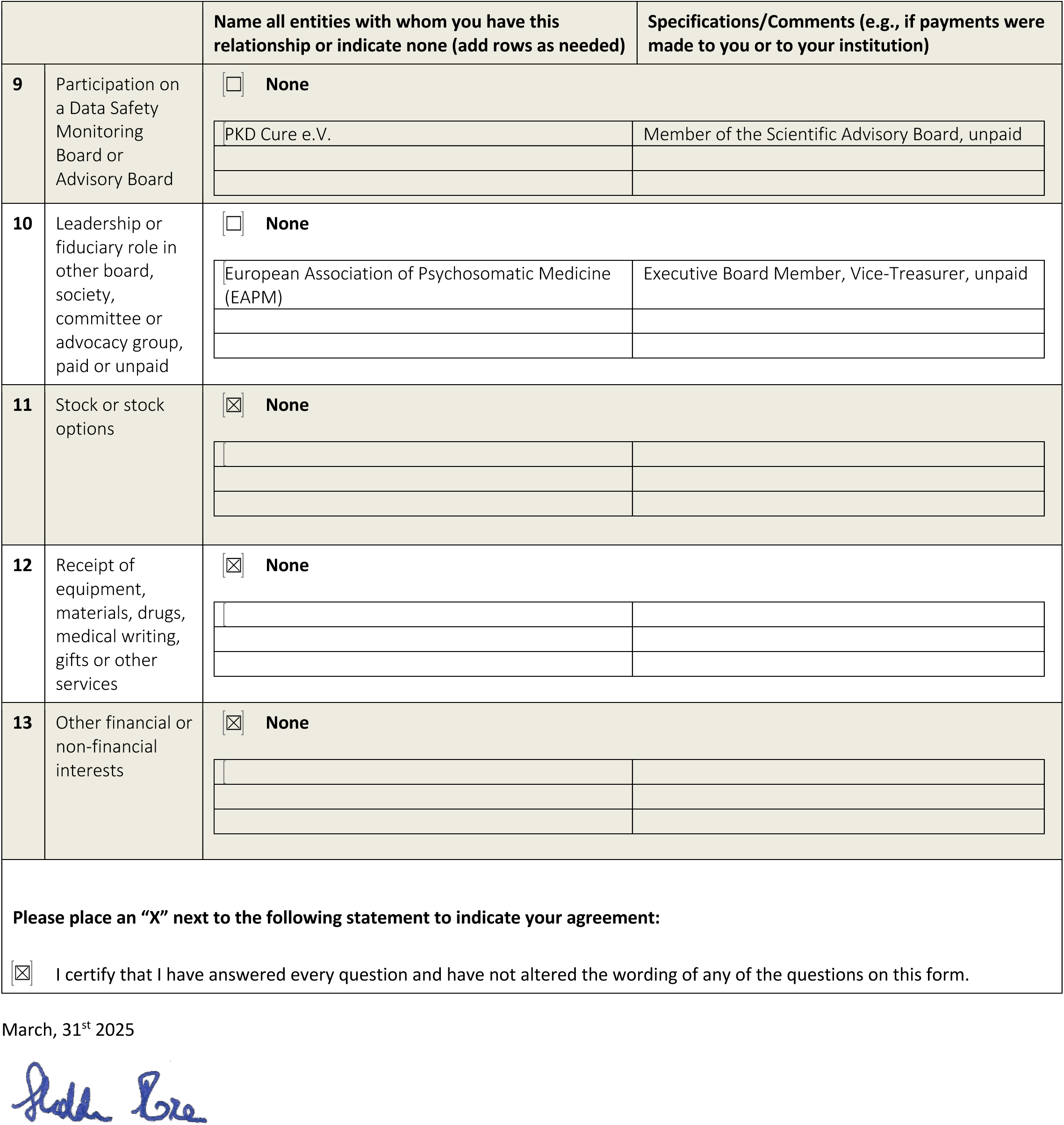

